# Causes and consequences of vascular dementia across the life course: Evidence from a UK Biobank phenome-wide and Mendelian randomization study

**DOI:** 10.1101/2025.06.23.25330106

**Authors:** Phazha Bothongo, Victoria Taylor-Bateman, Roxanna Korologou-Linden, Neil M Davies, Emma Hart, Patrick G Kehoe, Yoav Ben-Shlomo, Emma L Anderson

**Author notes:** Corresponding Author Phazha Bothongo, Address: Division of Psychiatry, University College London (UCL), 4^th^ Floor, Maple House, 149 Tottenham Court Road, London, W1T 7NF.

## Abstract

**Background:** Vascular dementia (VaD) is the second most common cause of dementia, yet its risk factors and biological mechanisms remain poorly understood.

**Objective:** To identify causes and consequences of VaD by developing a polygenic risk score (PRS) for VaD and conducting a phenome-wide association study (PheWAS), followed by Mendelian randomization (MR) analyses using data from the UK Biobank.

**Methods:** Using data from 334,758 UK Biobank participants, we first constructed a VaD PRS based on the most recent genome-wide association study (GWAS). We then performed an age-stratified PheWAS (39–53, 53–62, 62–72 years), examining 9,319 phenotypes associated with the VaD PRS. We followed up PheWAS hits with two-sample MR to evaluate causal relationships with VaD risk.

**Results:** Our PheWAS revealed age-dependent associations, with many relationships strengthening as age increased. Associations were found with vascular and Alzheimer’s dementias; cerebrovascular traits such as white matter hyperintensities (WMH), stroke, and intracerebral haemorrhage; adverse lipid profiles; elevated systolic blood pressure and glucose levels; reduced brain volumes (subcortical and hippocampal); and poorer cognitive function. The VaD PRS was also associated with higher risk of depression, Parkinson’s disease, neuroinflammatory disorders, and decreased basal metabolic rate and fat-free mass. MR analyses supported causal effects for WMH (OR: 1.83, 95% CI: 1.39–2.40), depression (1.25, 1.02–1.54), lipid traits (e.g., apolipoprotein B/A1 ratio: 1.31, 1.06–1.62), HbA1c (1.14, 1.02–1.28), and diastolic (1.03, 1.01–1.04) and systolic (1.01, 1.01–1.02) blood pressure. Protective factors included years of schooling (0.76, 0.64–0.90), apolipoprotein A (0.74, 0.59–0.92), fat-free mass (0.84, 0.71–0.99), and basal metabolic rate (0.82, 0.69–0.97).

**Conclusions:** Our findings highlight the central role of cardiometabolic and educational factors in the development of vascular dementia. Several modifiable risk factors—particularly blood pressure, glucose regulation, lipid levels, and years of schooling—showed evidence of causal effects on VaD risk. Age-stratified results suggest that early intervention, ideally from midlife, may offer the greatest preventive benefit by mitigating the progressive accumulation of vascular damage contributing to dementia risk.

## Introduction

Dementia is the leading non-communicable cause of death in the UK, yet it remains without effective preventive or disease-modifying treatments (1). While Alzheimer’s disease (AD) is widely studied, vascular dementia (VaD) has seen far less funding and research attention, despite cerebrovascular pathology being present in up to 80% of all dementia cases (2–4). This research gap hinders our understanding of VaD’s distinct biological mechanisms and limits the development of tailored interventions for both prevention and treatment.

VaD is a progressive neurocognitive disorder caused by impaired cerebral blood flow, resulting in brain injury (5,6). Prevention strategies primarily target modifiable cardiovascular risk factors such as cholesterol and blood pressure; however, evidence supporting their causal role in VaD remains limited due to challenges in early identification and diagnosis of VaD, particularly in the absence of reliable biomarkers. Most previous studies examining VaD risk factors have also been observational, making it difficult to establish causality or determine the directionality of effects due to the risk of confounding and reverse causation bias. Genome-wide association studies (GWAS) of vascular dementia have been limited in size. Currently, only the apolipoprotein E gene (*APOE*) has been robustly associated with VaD risk in the largest GWAS to date (3,892 European cases and 466,606 controls) (7). This is in stark contrast to the 75 genetic loci which have been associated with Alzheimer’s disease risk in the largest GWAS to date with over 111,326 European clinical and ‘proxy’ Alzheimer’s cases and 677,663 controls (8).

Large-scale genetic resources from large, longitudinally assessed cohorts such as the UK Biobank provide new opportunities to overcome these challenges. Phenome-wide association studies (PheWAS) can systematically evaluate associations between genetic liability for a disease and a wide array of phenotypic traits across organ systems and life stages (9–11). When combined with Mendelian randomization (MR), a method that utilises genetic variants as instruments to infer causality, this approach enables stronger inferences about both the causes and consequences of disease.

In this study, we apply a two-stage approach to investigate the life-course phenotypic manifestations of genetic liability to vascular dementia and to identify novel causal risk factors. First, we generate a VaD polygenic risk score (PRS) using the most recent VaD GWAS and conduct an age-stratified PheWAS across over 9,000 traits in the UK Biobank. Second, we apply two-sample MR to test for causal relationships with VaD. This combined approach enables the identification of both early biomarkers and modifiable risk factors that influence disease risk, providing insights into the timing and mechanisms of VaD development.

## Materials and Methods

### Study population

The UK Biobank is a population-based cohort study of approximately 500,000 individuals aged 40-69 years at recruitment (2006-2010), with comprehensive data on demographics, lifestyle, physical measurements, and health outcomes (12). From our initial 487,038 genotyped participants (full details in supplement), we excluded individuals with discrepant genetic/reported sex (n=367), sex chromosome aneuploidy (n=651), non-white British ancestry (n=78,320), third-degree or closer relatedness (n=81,788), and withdrawn consent (n=174). The final sample comprised 334,759 participants for analysis. A participant flow diagram is presented in Supplementary Figure 1.

### Study Design

We conducted a two-stage analysis to investigate age-dependent associations and potential causal relationships for VaD. First, we generated a PRS using the most recent GWAS of clinically diagnosed VaD and conducted a PheWAS across 9,319 traits, stratified by age tertiles. Traits included medical history, lifestyle factors, physical measures, cognitive tests, blood biomarkers, and brain imaging variables (Supplementary Table 1). We excluded administrative variables (e.g. assessment centre ID, timestamps, quality control flags), genetic data (as it was the exposure), sparse variables (<500 participants), and complex data types requiring non-standard analysis (Supplementary Table 2). In the second stage, we applied two-sample Mendelian randomization (MR) to all PheWAS hits to assess whether they represented potential causal risk factors, consequences of VaD genetic liability, or early disease biomarkers. The overarching research questions addressed by the PheWAS and MR analyses are summarised in Figure 1.

**Figure 1.**
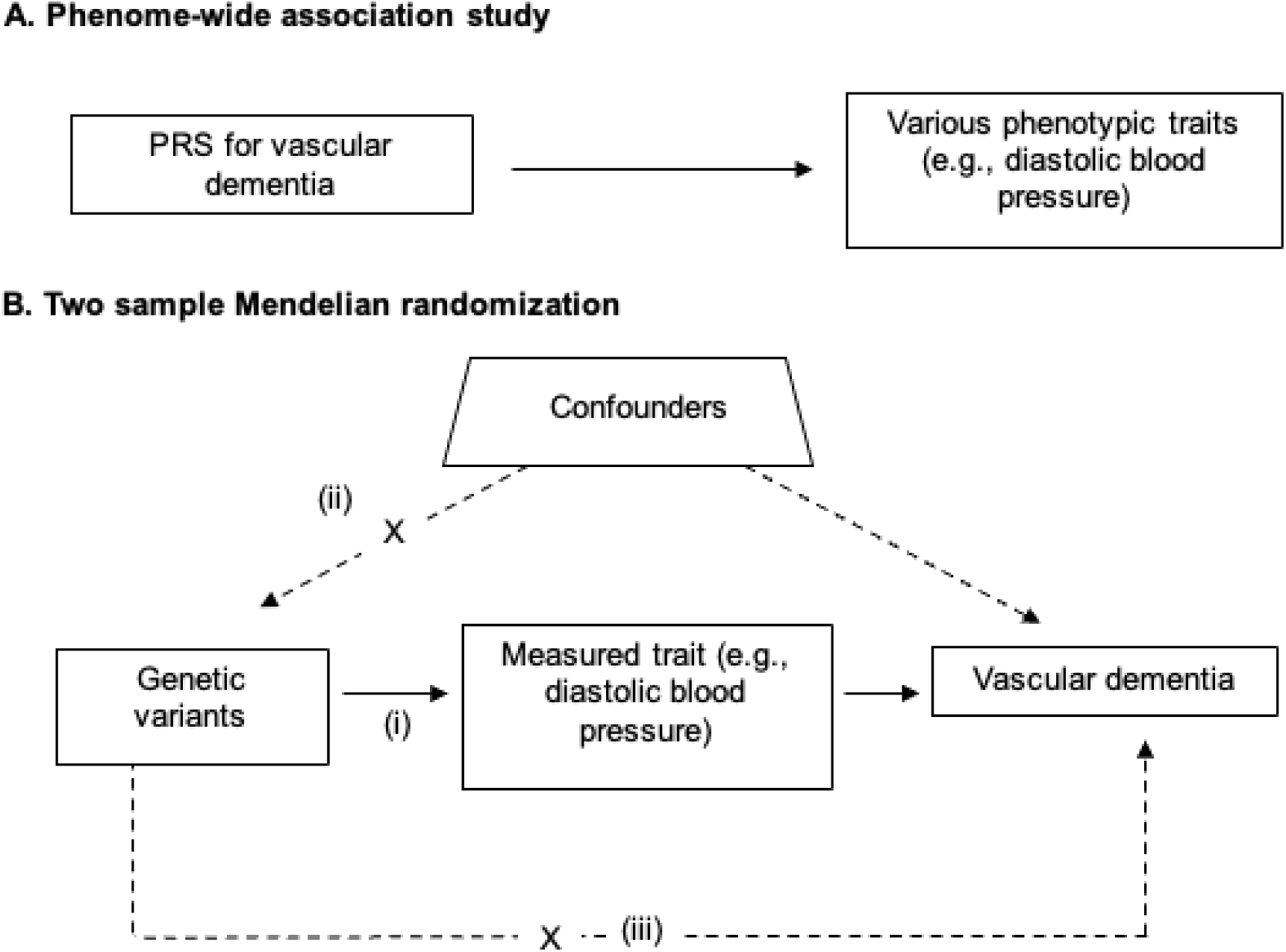
Study design for the phenome-wide association study of vascular dementia genetic liability, and follow-up Mendelian randomization of identified. Diagram (A) illustrates the study design for performing a phenome-wide association study (PheWAS), while diagram (B) depicts the approach for Mendelian randomization. In (A), the polygenic risk score (PRS) for vascular dementia (VaD) could influence a trait, such as diastolic blood pressure, either through a causal relationship or via alternative pathways unrelated to VaD (i.e., pleiotropy). Diagram (B) outlines the follow-up Mendelian randomization analysis aimed at determining whether the phenotypic associations are causal and determining direction.

This tests the hypothesis that a trait (e.g., diastolic blood pressure) causes increased risk of VaD. This relies on three key assumptions: (i) the single nucleotide polymorphisms (SNPs) used as instruments must be strongly associated with the trait (relevance); (ii) there must be no confounders of the SNP-outcome relationship association (independence); and (iii) the SNPs must influence the outcome exclusively through the trait of interest (exclusion restriction). X denotes that the assumption is that these pathways should not exist for MR to be valid.

## Statistical Analysis

### Generating Polygenic Risk Scores

For the forward PheWAS analysis (Figure 1A), we constructed a standardised, weighted PRS for clinically diagnosed VaD using all eligible UK Biobank participants (Supplementary Figure 1). The PRS included nine independent genetic variants associated with VaD at a suggestive significance threshold (P < 1 × 10^-6^), four of which met genome-wide significance (P < 5 × 10^-8^). These variants were identified through a meta-analysis combining two VaD GWAS datasets: (i) MEGAVCID (3,892 cases, 466,606 controls) (7) and (ii) FinnGen (3,116 cases, 433,066 controls) (13). The nine variants and their mapped genes are listed in Supplementary Table 3. The strongest association was observed for *APOE* (14), followed by genome-wide significant loci at *NECTIN2*, *APOC4*, and *APOC2*. PRSs were calculated by multiplying each participant’s allele count by the corresponding GWAS effect size and summing across all variants. Because only four variants reached genome-wide significance, we included suggestive variants to increase statistical power. However, this approach could potentially violate instrumental variable assumptions (see Figure 1). To assess the robustness of our findings, we conducted a sensitivity analysis using only the four genome-wide significant variants (Supplementary Table 4). Full details of this analysis are provided in the ‘Sensitivity Analysis’ section and Supplementary Methods.

### Phenome-wide association study

Given the late-life onset of VaD and the relatively young age of UK Biobank cohort, we stratified participants into three equally sized age tertiles (n=111,586 per tertile): tertile 1 (age 39-53 years, mean=47.2±3.8 years, 55% female); tertile 2 (age 53-62 years, mean=58.0±2.4 years, 55% female); and tertile 3 (age 62-72 years, mean=65.3±2.3 years, 51% female). This stratification enabled identification of potential early biomarkers in preclinical VaD, potential causal risk factors, and age-specific effects.

Using the MR-PHESANT R package (15), we regressed each UK Biobank variable on the VaD PRS within each age tertile, and then within the entire sample, see Supplementary Tables 5-8. The methodology behind MR-PHESANT’s automated rule-based approach has been detailed in previous publications (15,16). To reduce standard errors, all models were adjusted for age, sex, assessment centre and genotype batch. We also adjusted for the first ten genetic principal components to minimise bias due to population stratification. Multiple testing was addressed using the false discovery rate (FDR) correction at 5%. For continuous outcomes, the MR-PHESANT applies an inverse rank-normal transformation. Thus, in this case, results represent the effect on inverse rank-normal transformed outcomes, per standard deviation (SD) increase in the PRS. For binary outcomes, results are presented as log-odds ratios.

### Follow-up Mendelian Randomization Analysis

We selected phenotypes for two-sample MR follow-up based on two criteria: (1) PheWAS associations with an FDR-corrected P value ≤ 0.05 and (2) an *a priori* defined list of hypothesised risk factors for VaD from previous literature (17–20). This predefined list included educational attainment (years of schooling), cognitive function (fluid intelligence), systolic and diastolic blood pressure, obesity, hearing loss, smoking, depression, physical inactivity, social isolation, type 1 and 2 diabetes, excessive alcohol consumption, air pollution, body mass index (BMI), glycated haemoglobin (HbA1c), high intake of sugar-sweetened beverages, total cholesterol, low density lipoprotein cholesterol (LDL), high density lipoprotein cholesterol (HDL). We also examined white matter hyperintensity volume (WMH) as a biomarker of VaD pathology. To ensure robust instrumental variable analysis, we excluded phenotypes with fewer than 5 available genetic variants as instruments or with F statistics below 10, minimising weak instrument bias (Supplementary Table 9-10) (21). For the remaining phenotypes, we implemented two-sample MR using summary statistics from GWAS which were independent of the UK Biobank, where available (Supplementary Table 11). Where sample overlap occurred with the UK Biobank, sensitivity analyses were performed with MRlap to examine potential bias (further detail in ‘Sensitivity analyses). Our primary MR method was inverse variance weighted (IVW) regression, which provides the most precise estimates when all genetic variants are valid instruments and assumes no horizontal pleiotropy. MR results are presented as odds ratios with 95% confidence intervals for binary outcomes and beta coefficients with standard errors for continuous outcomes (Supplementary Tables 12-14).

### Sensitivity analyses

Firstly, given that we included five VaD genetic variants with only suggestive evidence in the PRS (*P* < 1 x 10^-6^), we replicated the PheWAS using only the four genome-wide significant variants (*P* < 5 x 10^-8^) to examine whether results were comparable (Supplementary Tables 15-17). Secondly, we tested for linear trends in effect estimates across age tertiles using meta-regression (22) for previously implicated dementia risk factors, with age tertile as the independent variable and trait beta coefficients as dependent variables (full results in Supplementary Table 18). Thirdly, we examined potential bias due to horizontal pleiotropy with MR Egger and weighted median estimators. We also assessed heterogeneity using Cochran’s Q statistics for significant estimates (P ≤ 0.1) (Supplementary Table 19). Lastly, to address potential for bias from sample overlap between exposure and outcome datasets in the follow-up two-sample MR (23), we implemented MRlap for all analyses (24), which is robust to sample overlap, weak instrument bias, and winner’s curse (24).

## Results

### PRS of vascular dementia

The distribution of the VaD PRS in VaD cases and controls is shown in Supplemental Figure 2. A SD increase in the VaD PRS was strongly associated with vascular dementia (OR= 1.52, 95% CI: 1.46 to 1.59, R^2^=0.08), Alzheimer’s disease (OR= 1.86, 95% CI: 1.81 to 1.91, R^2^=0.05) and all-cause dementia (OR =1.63, 95% CI: 1.60 to 1.66, R^2^=0.03). The VaD PRS was also associated with WMH volume in the oldest tertile (beta= 0.03, 95% CI: 0.01 to 0.040, R^2^=0.0003).

### PheWAS of Vascular Dementia Polygenic Risk Score

Full PheWAS results for all age tertiles and the entire cohort are presented in Supplemental Tables 5-8.

#### Associations with Previously Implicated Dementia Risk Factors

The VaD PRS was associated with increased total and LDL cholesterol, SBP, WMH volume, moderate and vigorous physical activity levels, and depressive episodes (Figure 2). It was also associated with lower HDL cholesterol, HbA1c, BMI, DBP, smoking, alcohol consumption, obesity and type 2 diabetes risk, fluid intelligence scores and exposure to air pollution (Figure 2). Effect estimates were broadly consistent across age groups, with confidence intervals largely overlapping, except for total cholesterol (P trend = 4.11×10^-20^) and LDL cholesterol (β trend = −0.024, P trend = 9.27×10^-19^), where associations were largest in the middle age tertile (Figure 2).

**Figure 2.**
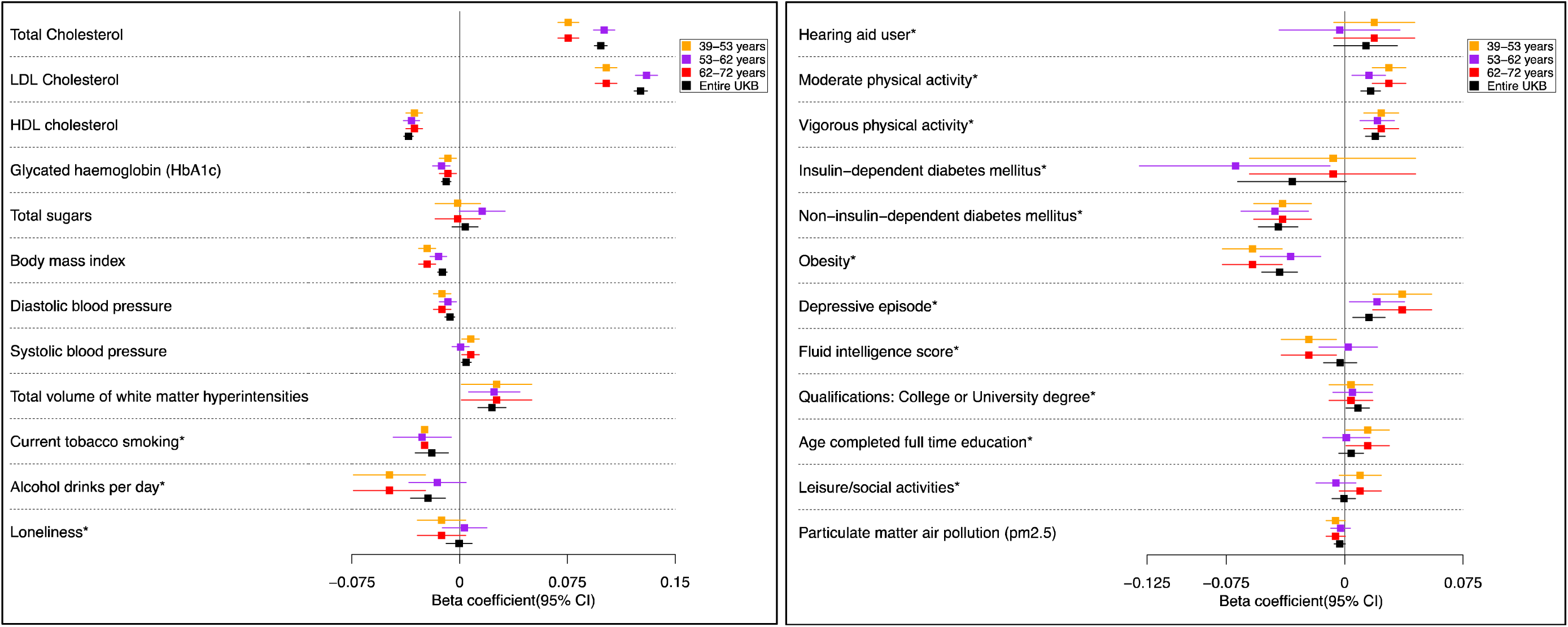
Forest plots showing effect estimates for the association between vascular dementia polygenic risk score and previously implicated risk factors of dementia, by age tertile. Legends in each graph indicate age tertiles and entire sample. Effect estimates represent an SD change in the phenotype per 1 unit increase in the standardized polygenic risk score for vascular dementia. Error bars represent 95% confidence intervals. Each tertile consists of 111,656 participants and the exact sample size for each phenotype are in the Supplementary Data file. Effect estimates were derived from linear regression models and are in standard deviations. *Effect estimates were derived from binary logistic models; effect estimates are on the log odds scale.

#### Associations with neurological and cognitive measures

The VaD PRS was positively associated with variables capturing family history of dementia, with most effects increasing in magnitude with age (Figure 3). The VaD PRS was also consistently associated with poorer performance on various cognitive domain tests, particularly in the middle and oldest age tertiles, and a greater risk of Parkinson’s disease, including secondary and all-cause Parkinson’s disease, epilepsy and encephalitis in the oldest age tertile.

**Figure 3.**
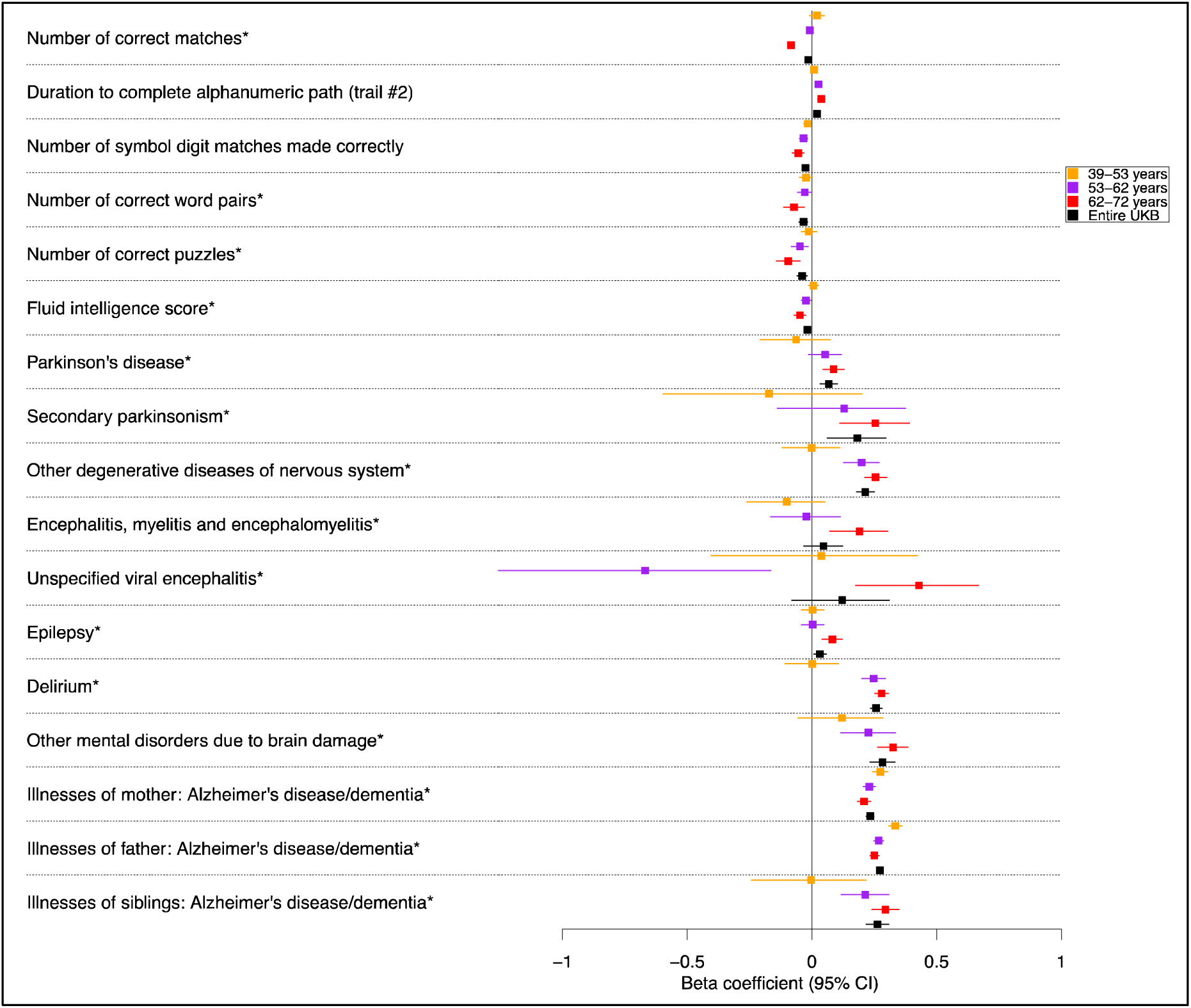
Forest plots showing effect estimates for the association between vascular dementia polygenic risk score and neurological conditions and cognitive function tests, by age tertile. Legends in each graph indicate age tertiles and entire sample. Effect estimates represent an SD change in the phenotype per 1 unit increase in the standardized polygenic risk score for vascular dementia. Error bars represent 95% confidence intervals. Each tertile consists of 111,656 participants and the exact sample size for each phenotype are in the Supplementary Data file. Effect estimates were derived from linear regression models and are in standard deviations. *Effect estimates were derived from binary logistic models; effect estimates are on the log odds scale.

#### Associations with brain imaging measures

Associations of the VaD PRS and brain imaging parameters are shown in Supplementary Figure 3. A higher VaD PRS was negatively associated with grey matter volume in key subcortical and limbic regions (amygdala, ventral striatum, hippocampus) in the two oldest age tertiles, and with hippocampal volume, fornix integrity, and precuneus grey-white contrast in all age groups.

#### Associations with cardiometabolic traits

The VaD PRS was positively associated with cerebrovascular diseases, with magnitudes of effects progressively strengthening with age (Figure 4). It was also associated with increased risk of lipoprotein metabolism disorders, angina, acute myocardial infarction, heart attack, ischemic heart disease, intracerebral haemorrhage, hypotension and stroke, particularly in the middle and oldest age tertiles. The VaD PRS was negatively associated with basal metabolic rate in the middle and oldest age tertiles.

**Figure 4.**
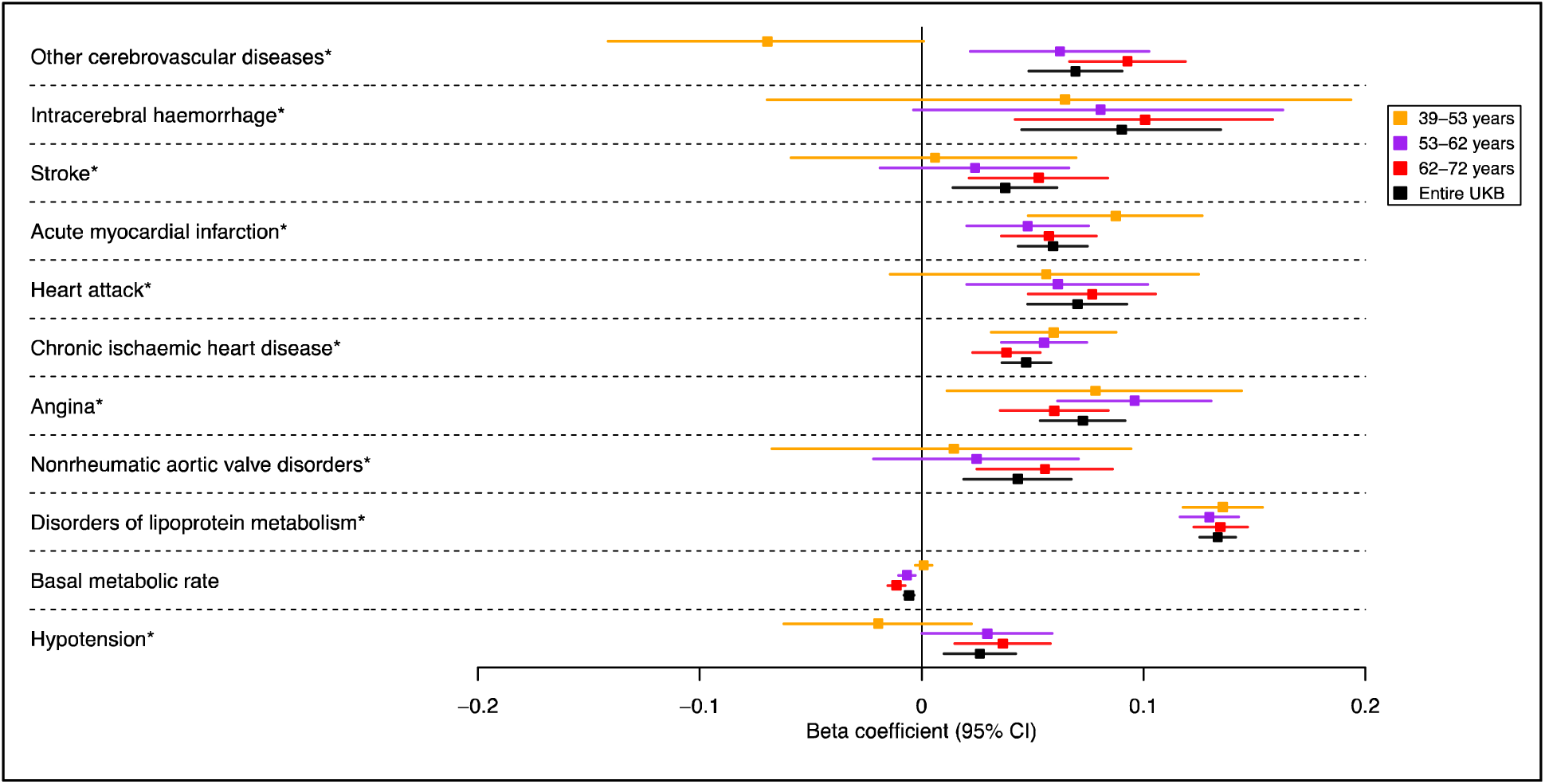
Forest plots showing effect estimates for the association between vascular dementia polygenic risk score and cardiovascular & metabolic medical history, by age tertile. Legends in each graph indicate age tertiles and entire sample. Effect estimates represent an SD change in the phenotype per 1 unit increase in the standardized polygenic risk score for vascular dementia. Error bars represent 95% confidence intervals. Each tertile consists of 111,656 participants and the exact sample size for each phenotype are in the Supplementary Data file. Effect estimates were derived from linear regression models and are in standard deviations. *Effect estimates were derived from binary logistic models; effect estimates are on the log odds scale.

#### Associations with blood parameters and biological measures

The VaD PRS was consistently associated with lower levels of several blood parameters (Supplemental Figure 4-5) including red blood cell count and distribution width, plateletcrit and platelet count, reticulocyte volume, haematocrit percentage, haemoglobin concentration and monocyte count. The VaD PRS was also positively associated with several blood-based measures of lipid-related phenotypes (ApoB, LDL, IDL, VLDL, fatty acids, and triglycerides), with magnitudes of effect typically decreasing with age (figure 5). A higher VaD PRS also positively associated with markers related to liver function and metabolism (total bilirubin, 3-Hydroxybutyrate), growth regulation (IGF-1), and fatty acid composition and unsaturation (linoleic acid to fatty acids percentage, degree of unsaturation, docosahexaenoic acids, omega-3 fatty acids), with similar effect sizes observed across all age groups. Conversely, the VaD PRS was negatively associated with ApoA, ApoA1, HDL, kidney function markers, and several inflammatory markers such as C-reactive protein and monocyte count, with comparable magnitudes of effects across age groups.

**Figure 5.**
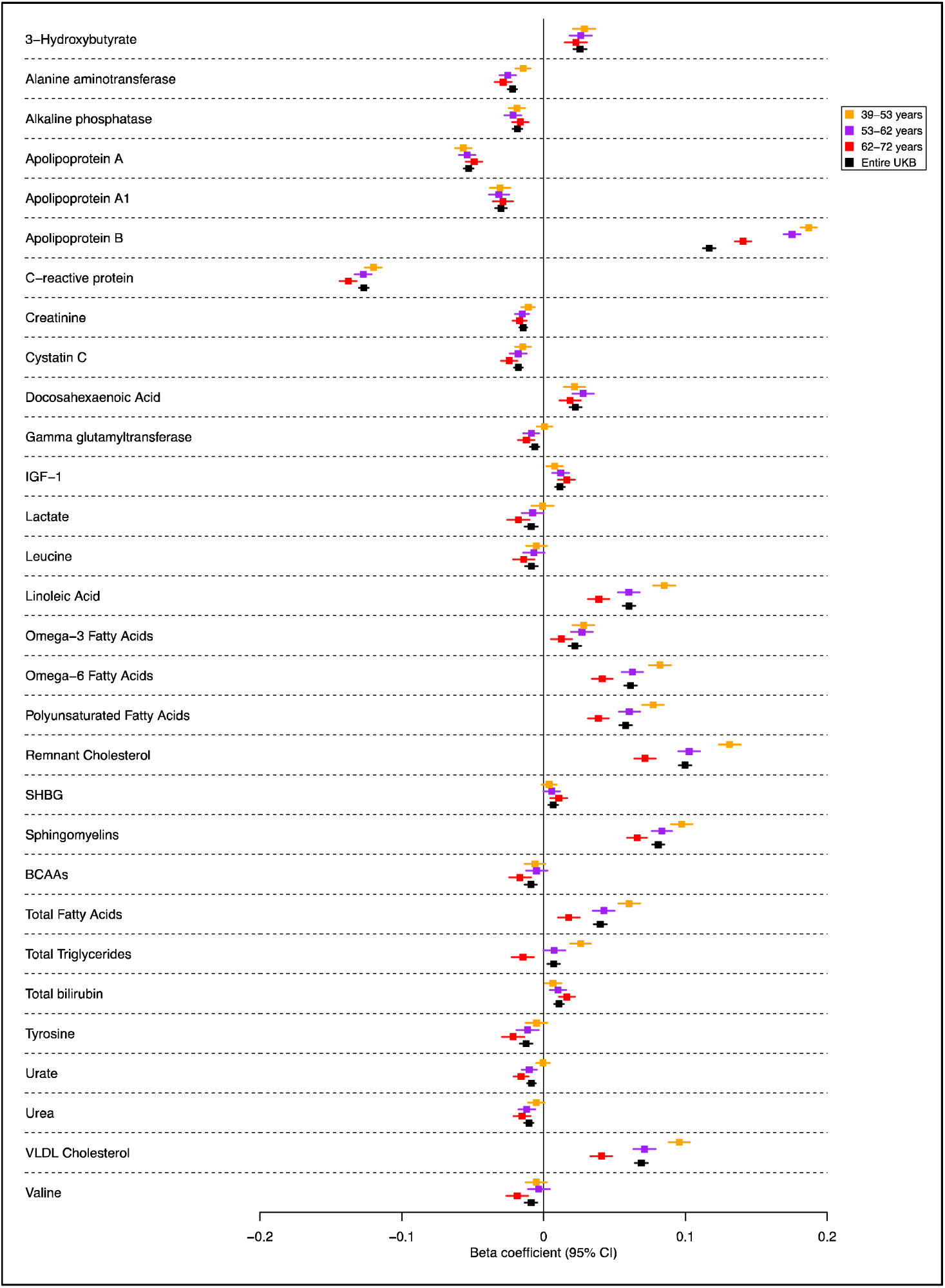
Forest plots showing effect estimates for the association between vascular dementia polygenic risk score and biological measures from blood assay results, by age tertile. Legends in each graph indicate age tertiles and entire sample. Effect estimates represent an SD change in the phenotype per 1 unit increase in the standardized polygenic risk score for vascular dementia. Error bars represent 95% confidence intervals. Each tertile consists of 111,656 participants and the exact sample size for each phenotype are in the Supplementary Data file. Effect estimates were derived from linear regression models and are in standard deviations.

#### Associations with medication use and immune related measures

A higher VaD PRS positively associated with Aricept (donepezil) use, which is a treatment for symptoms of Alzheimer’s disease, and negatively associated with Buscopan use; a treatment for painful stomach cramps due to menstruation or irritable bowel syndrome, in the oldest tertile (Supplementary Figure 6). The VaD PRS also positively associated with lipid-lowering medication use, aspirin use, and omega-3 supplementation in all age groups. A higher VaD PRS was positively associated with neuroinflammatory disorders (unspecified viral encephalitis, encephalitis, myelitis and encephalomyelitis), and negatively associated with several infectious diseases (chickenpox, mumps, measles) in the oldest tertile (Supplementary Figure 7).

#### Other associations

Several other associations were identified between the VaD PRS and downstream measures – for example, negative associations were seen with several respiratory parameters (including peak expiratory flow, diaphragmatic hernia, wheezing in chest, interstitial pulmonary disease), hypersensitivity pneumonitis, several body composition measures (generally the VaD PRS was associated with lower total body fat on various indicators of adiposity), longer hearing test response, and several eye parameters (including a positive associations with spherical power, and average mean spherical equivalent) (see ‘additional results’ section of the online supplement and Supplementary Figure 8-16).

### Two-sample Mendelian randomization follow-up of risk factors for vascular dementia

Several traits showed evidence of being a causal risk factor for VaD (Figure 6, full results in Supplementary Tables 11-13 and 17). The following phenotypes were positively associated with the risk of clinical diagnosis of VaD (IVW *P* value ≤ 0.05): depression, schizophrenia, higher diastolic and systolic blood pressure, greater white matter hyperintensity volume, total lipids, high HbA1c, phospholipids, and c-reactive protein. There was also suggestive evidence (IVW *P* value > 0.05 & ≤ 0.10) that the following phenotypes are positively associated with VaD risk: coronary heart/artery disease, higher BMI, increased oily fish and salad/raw vegetable intake, cholesteryl esters, VLDL, sphingomyelins, gamma-glutamyl transferase, and higher haemoglobin concentration.

**Figure 6.**
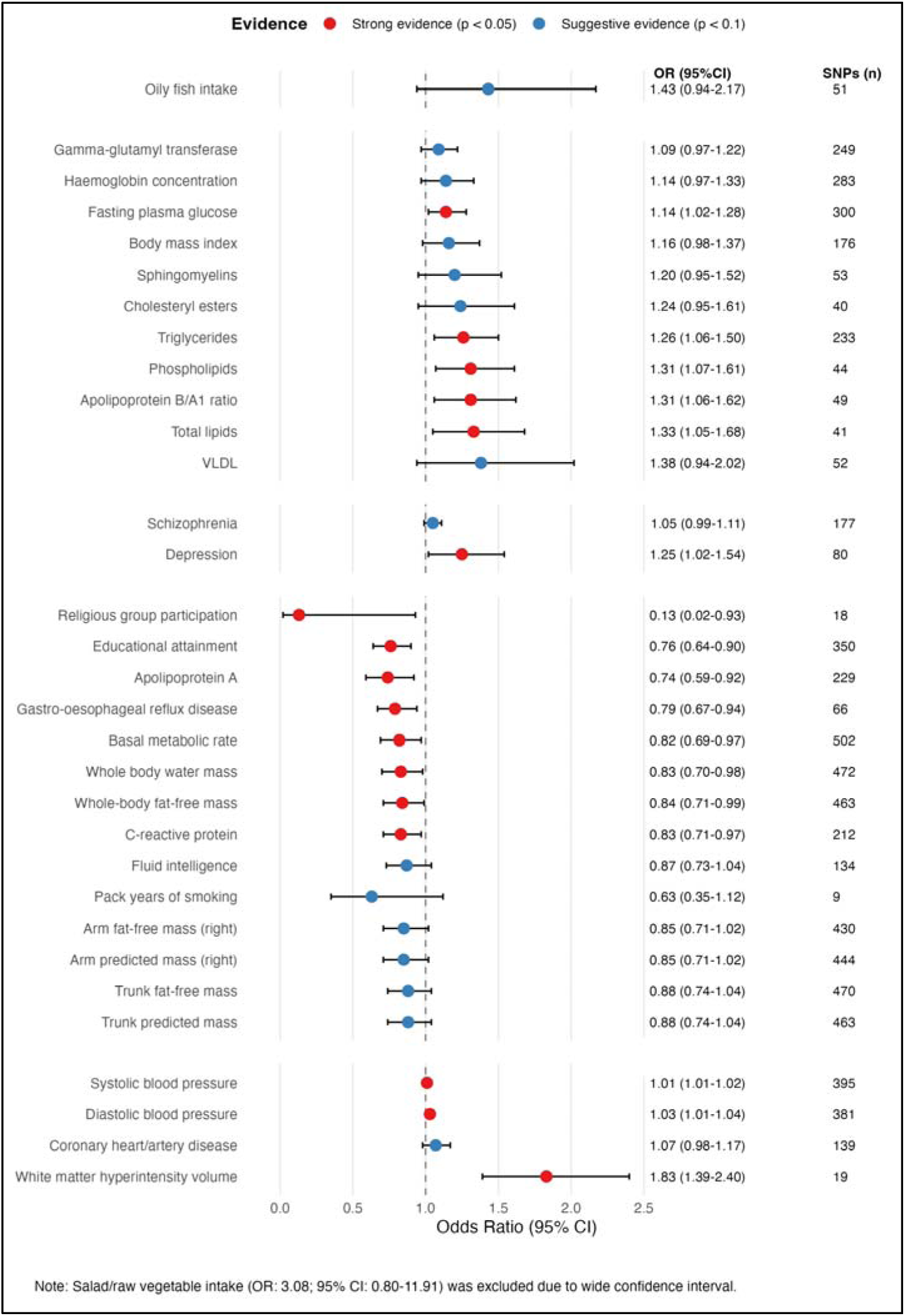
Effect estimates with 95% confidence intervals examining the causal association between PheWAS hits and vascular dementia, using Mendelian Randomization

The following phenotypes were negatively associated with the risk of VaD (IVW *P* value ≤ 0.05): gastro-oesophageal reflux disease, higher educational attainment, higher basal metabolic rate, higher whole body water mass, whole-body fat-free mass, religious group participation, triglycerides, apolipoprotein A, and apolipoprotein B/A1 ratio. There was also suggestive evidence (IVW *P* value > 0.05 & ≤ 0.10) that the following phenotypes were negatively associated with VaD risk: higher fluid intelligence, years of smoking, arm fat-free mass (right), arm predicted mass (both right and left), trunk fat-free mass, and trunk predicted mass.

### Sensitivity analyses

Overall, PheWAS results were very comparable when using a VaD PRS which only contained the four genome-wide significant VaD variants and no suggestively associated variants; 78% (n=435) of the original 558 PheWAS hits detected in the main analysis were also detected when using this restricted PRS, and results of the two PheWAS were highly correlated (Supplementary Figure 17-19 for correlation plots within each age tertile). Follow-up MR indicated some horizontal pleiotropy across several traits including blood assays, anthropometric and metabolic traits (e.g., trunk/arm mass, basal metabolic rate), cardiovascular traits (e.g., coronary heart disease, blood pressure), and lifestyle factors (e.g., years smoking; see Supplemental Table 12). MR results were consistent when using MRlap to correct for sample overlap bias. Where bias was detected, IVW effect sizes were typically underestimated (e.g., schizophrenia, depression, WMH, BMI; see Supplemental Table 12 and Supplementary Figure 20). MRlap also revealed causal effects for several phenotypes that were not detected by the FDR-corrected IVW estimates, including positive associations with Parkinson’s disease and vitamin D, several cognitive tests (e.g., positive associations for trail making and reaction time, negative associations for digit substitution), metabolic and body composition factors (e.g., positive associations for fat percentage and obesity, negative associations for impedance), biological measures (e.g., positive associations for alanine aminotransferase, degree of unsaturation, and negative associations for apolipoprotein B, chylomicrons, haematocrit, IGF-1, platelet crit, and red blood cell count), and lifestyle traits (e.g., positive associations for physical activity and non-oily fish intake; Supplemental Table 13).

## Discussion

We conducted a hypothesis-free PheWAS to examine how the VaD PRS affects the phenome across the life course. Associations were evident across all age groups, with many strengthening in older adults (62-72 years), but early phenotypic manifestations were also observed in younger participants (39-53 years). Two-sample MR analysis enabled us to begin disentangling potential causes from consequences of VaD.

Our findings support effective and early cardiovascular risk factor management for VaD prevention, with potential benefits across the spectrum of cerebrovascular cognitive impairment, consistent with existing clinical guidelines (25). However, the optimal timing of intervention is complex. Several factors must be considered, including the latency between exposure and disease onset, the potential for reversing pathological changes at different life stages, and the cost-effectiveness of interventions across age groups. Our age-stratified results suggest that while early intervention is likely to yield the greatest benefits (given the progressive accumulation of cerebrovascular damage), different risk factors may be more or less influential at different ages. This highlights the importance of age-specific prevention strategies rather than a ‘one-size-fits-all’ approach to all risk factors.

In our PheWAS, the VaD PRS was positively associated with cerebrovascular diseases (such as intracerebral haemorrhage) and systolic blood pressure (SBP) but negatively associated with diastolic blood pressure (DBP) and body mass index (BMI) across all age tertiles. The inverse association between the VaD PRS and DBP may reflect the complex physiology of diastolic pressure. DBP is influenced by both the vasoconstrictor tone of arterioles (where smaller vessel radius increases DBP) and arterial compliance (where more elastic vessels also support higher DBP) (26–28). In stiffer, less compliant arteries— common in aging—SBP tends to increase, while DBP may paradoxically decrease (29). This widening of pulse pressure (high SBP, low DBP) is a hallmark of vascular aging and suggests that lower DBP is not necessarily protective; instead, it may be a marker of arterial stiffness; a likely risk factor for vascular dementia (29). Thus, DBP may not be a reliable standalone biomarker of VaD risk. This is consistent with findings from the Framingham Heart Study, which identified SBP as a stronger predictor of cardiovascular outcomes than DBP (30). Supporting this, our MR analysis provided strong evidence that both higher SBP and DBP increase VaD risk. This suggests that maintaining well-controlled blood pressure— especially earlier in life, before vascular stiffening occurs—may reduce future VaD risk. We also observed associations between the VaD PRS and hypotension, particularly in the middle and oldest age tertiles. It is plausible that this reflects a failure of the brain to maintain adequate cerebral perfusion through autoregulatory mechanisms. In older adults, age-related vascular stiffening and arteriolar degeneration impair cerebral autoregulation, such that even modest drops in systemic blood pressure can reduce cerebral blood flow, especially in white matter regions vulnerable to hypoperfusion (31–33). Cohort studies including the Rotterdam Study and the Kungsholmen Project have shown that low diastolic blood pressure is associated with increased risk of cognitive decline and dementia, likely due to impaired perfusion of the aging brain (34,35). These findings support the hypothesis that elevated blood pressure may in some cases be a compensatory response to cerebral hypoperfusion, and that failure to maintain this response (resulting in hypotension) could contribute to the progression of vascular neurodegeneration.

Our study identified lipid metabolism as a key contributor to vascular brain health. The VaD PRS was positively associated with multiple lipid-related phenotypes, and the PheWAS revealed that individuals with higher genetic liability to VaD had lower HDL, higher LDL and total cholesterol levels, as well as increased use of lipid-lowering medications. However, whilst there was some evidence of total lipids as an aggregate measure on VaD risk, our follow-up MR analyses did not support a causal effect of HDL, LDL or total cholesterol specifically on VaD risk. This suggests the PheWAS associations may reflect shared genetic pathways or reverse causation, where early vascular changes influence lipid levels, rather than direct causality. This is also in line with a recent drug target MR study which found very little evidence to suggest that lipid-lowering medications affect risk of VaD diagnoses or it’s neuroimaging biomarkers (36). Overall, these findings underscore the importance of examining specific lipid fractions in vascular dementia research, and also caution against overinterpreting observational associations.

Our analysis confirmed a substantial association between WMH volume and a clinical diagnosis of VaD (83% increased risk of diagnosis per SD increase in WMH), supporting WMH as a key biomarker of underlying vascular pathology as a driver of cognitive impairment. This finding aligns with longitudinal imaging studies such as the Framingham Offspring Study by Debette et al. (2010), which demonstrated that WMH burden predicted subsequent cognitive decline, dementia, and mortality (37,38). Associations between the VaD PRS and neurological disorders, including Parkinson’s disease, cognitive function measures, and brain imaging phenotypes, were also evident in our analysis across all age tertiles. The observed association with Parkinson’s disease may reflect vascular parkinsonism being misdiagnosed as idiopathic Parkinson’s disease, a known clinical challenge (39,40). We also found that higher genetic liability to VaD was associated with reduced grey matter volume in key subcortical regions, supporting evidence of early structural brain changes. These findings are consistent with the hypothesis that VaD risk is linked to both vascular and neurodegenerative pathology, and that genetic predisposition may manifest through measurable changes in brain morphology, long before clinical symptoms emerge (7,41,42).

Genetic liability to psychiatric disorders, such as schizophrenia and depression, was positively associated with VaD risk in our MR analysis, suggesting that either these conditions may directly contribute to disease development, or that there are potential shared pathophysiological mechanisms. In our PheWAS, a higher VaD PRS was also associated with increased hearing loss, which may reflect neurodegenerative changes affecting sensory processing. However, our MR analysis found little evidence to support a causal role of hearing loss in the development of VaD, suggesting it is more likely a consequence of accumulating cerebrovascular damage and its downstream effects on neural integrity, rather than a contributing cause of the disease.

Our PheWAS and MR analyses both supported the protective role of educational attainment in vascular brain health, echoing what has been previously observed for Alzheimer’s disease (43). Our findings lend support to both the brain (i.e. physical capacity) and cognitive (i.e. functional adaptability) reserve hypotheses in vascular dementia. Brain reserve may confer resilience through enhanced collateral circulation, improved blood flow regulation, and greater neural tolerance to reduced oxygen supply, while cognitive reserve may involve compensatory cognitive mechanisms that help maintain function despite accumulating underlying cerebrovascular damage (44–47). Social engagement, particularly religious group participation, also showed evidence of protective effects against VaD, aligning with previous evidence for social participation’s benefits for cognitive health as documented in Livingston et al.’s (2024) Lancet Commission report and Evans et al.’s (2019) meta-analysis, showing that social isolation increases dementia risk by approximately 50% (48,49). However, it is worth noting that confidence intervals for the religious group participation estimate in our study were very wide, suggesting a lack of power.

Lastly, we identified a novel protective association between gastro-oesophageal reflux disease (GERD) and VaD in the MR analysis. This finding could reflect several plausible mechanisms. Hormonal influences may play a role, as oestrogen and progesterone increase nitric oxide synthesis, a muscle relaxant which decreases smooth muscle tone of the lower esophageal sphincter, predisposing to gastro-oesophageal reflux (50). Recent meta-analyses from 2023 demonstrate that hormone therapy users had significantly increased GERD risk, with oestrogen alone contributing 41% higher odds and progesterone-only therapy 39% higher risk (51). Alternatively, individuals who use aspirin for cardiovascular protection are more at risk of GERD, and recent large cohort studies show that long-term low-dose aspirin demonstrates protective potential against both vascular dementia and Alzheimer’s disease, specifically in patients with coronary heart disease (52). This association warrants further investigation into shared biological pathways or potential medication effects. Such unexpected findings highlight the value of hypothesis-free approaches like PheWAS in generating new research directions.

It is worth noting that the direction of some associations identified in the PheWAS were contrary to what might be expected *a prior*i. For example, a higher VaD PRS was associated with lower HbA1c levels, type 2 diabetes risk, BMI, obesity, smoking and alcohol consumption, and higher physical activity. These associations could potentially reflect, at least in part, selection bias in the UK Biobank, where participants are known to be healthier than the general population and less likely to be obese, smoke, or drink alcohol regularly.

Supporting this, our two-sample MR analysis found little evidence that HbA1c, BMI, smoking or alcohol consumption is causally related to VaD risk. We did, however, find MRlap evidence to suggest higher physical activity levels increased VaD risk, and this has also been previously observed for Alzheimer’s disease (53). This could plausibly reflect survival bias whereby those who are more physically active, on average, live longer and are therefore more likely to receive a VaD diagnosis.

### Strengths and limitations

Our study leverages comprehensive phenotyping in the UK Biobank, age-stratified analysis to explore when associations emerge, and a two-stage design combining PheWAS with MR to examine both directionality and causality. The large, well-characterised sample offers exceptional statistical power to detect phenotypic manifestations of genetic VaD risk across the life course. Our systematic, hypothesis-free approach also reduces bias inherent to targeted analyses. Notably, the relatively young UK Biobank population allows assessment of preclinical risk patterns before diagnosis, reducing survival bias common in older cohorts.

There are several limitations to our study. Like all MR analyses, ours relies on core assumptions of relevance, independence, and exclusion. Relevance is supported by including only genome-wide significant SNPs with F-statistics >10. Independence is likely upheld through restriction to European ancestry to minimise bias due to population structure. Potential violations of the exclusion restriction were assessed using MR-Egger, weighted median/mode, and Cochran’s Q. Some traits showed evidence of horizontal pleiotropy, which we acknowledge in our interpretation (24,54) UK Biobank participants are healthier and more affluent than the general population, potentially limiting generalisability. Further replication in more diverse populations is needed. While PheWAS in non-European UKB subgroups is possible, no robust VaD PRS currently exists for non-European ancestries due to small case numbers (African ancestry: N=96 cases; Asian ancestry: N=115 cases; Hispanic ancestry: N=35 cases) (14). This limits the options for cross-ancestry analyses, especially given APOE’s known ancestry-specific effects on dementia and brain aging more broadly (55,56). Finally, the VaD GWAS used to construct the PRS was relatively small (7,008 cases vs 899,672 controls) compared to other dementia GWAS (e.g. Alzheimer’s disease: 111,326 cases) (8). To boost statistical power, we included both genome-wide significant and suggestive SNPs. While this has the potential to introduce some non-causal signals, results were consistent when restricted to genome-wide significant variants.

## Conclusions

Vascular dementia risk arises from diverse, age-dependent pathways. Our findings highlight the central role of cardiometabolic health, with robust evidence for several modifiable risk factors including elevated blood pressure, impaired glucose regulation, dyslipidaemia, as well as lower educational attainment. Our findings underscore the urgent need for more holistic, integrated, and multi-domain prevention strategies within health care systems, starting earlier in life, from around age 40 onwards. By clarifying *when* and *how* vascular risk factors converge on brain health, our study provides a roadmap for earlier and more targeted interventions to help reduce the growing global burden of vascular dementia.

## Supporting information

Supplementary Methods and Results

## Data Availability

Data are available from the UK Biobank (application ID: 123335) and can be accessed through the UK Biobank Access Management System (https://www.ukbiobank.ac.uk/enable-your-research/apply-for-access). GWAS summary statistics are available from MEGAVCID consortium and FinnGen. Analysis code is available upon reasonable request.

## Acknowledgements

The genetic and phenotypic information utilized in this research was accessed from the UK Biobank under application ID: 123335. We are also grateful for the collaboration of the MEGAVCID consortium who shared their VaD GWAS summary statistics which enabled this analysis to be performed.

