## Supplementary Methods and Results for "Causes and consequences of vascular dementia across the life course: Evidence from a UK Biobank phenome-wide and Mendelian randomization study"

### Supplementary Table of Contents

[Supplementary Figure 17. Scatterplot comparing all beta estimates from the original and adjusted vascular PheWAS analyses in the oldest tertile group. Plot compares beta estimates from the original PheWAS (blue dots), which included suggestive (p < 1x10^-6^) SNPs, and the adjusted PheWAS (red dots) where we only included genome-wide significant (p < 5x10^-8^) SNPs in our PRS. Significant associations (false discovery rate (FDR) < 5%) in the adjusted analysis are highlighted in green.	29](#_Toc200451556)

[Supplementary Figure 18. Scatterplot comparing all beta estimates from the original and adjusted vascular PheWAS analyses in the middle tertile group. Plot compares beta estimates from the original PheWAS (blue dots), which included suggestive (p < 1x10^-6^) SNPs, and the adjusted PheWAS (red dots) where we only included genome-wide significant (p < 5x10^-8^) SNPs in our PRS. Significant associations (false discovery rate (FDR) < 5%) in the adjusted analysis are highlighted in green.	31](#_Toc200451557)

[Supplementary Figure 19. Scatterplot comparing all beta estimates from the original and adjusted vascular PheWAS analyses in the youngest tertile group. Plot compares beta estimates from the original PheWAS (blue dots), which included suggestive (p < 1x10^-6^) SNPs, and the adjusted PheWAS (red dots) where we only included genome-wide significant (p < 5x10^-8^) SNPs in our PRS. Significant associations (false discovery rate (FDR) < 5%) in the adjusted analysis are highlighted in green.	32](#_Toc200451558)

[Supplementary Figure 20. Significant and suggestive association of vascular dementia polygenic risk score with the phenome, and estimated effect of each phenotype using Mendelian randomization. These findings showed evidence of association in the MR framework, following a correction for multiple testing using a strategy controlling for the false discovery rate. + and – indicate the direction of the coefficient for phenotypes associated with vascular dementia using two-sample MR. X represents associations that were consistent with the null. *Previously implicated risk factors of all-cause dementia.	34](#_Toc200451560)

### Supplementary Methods

#### Previously Implicated Risk Factors, with UK Biobank Equivalent Risk Factors

(a) (1)

1. **Less education:** Qualifications, Age completed full time education.
2. **Midlife hypertension**: Systolic blood pressure (automated reading), Diastolic blood pressure (automated reading).
3. **Obesity:** Date E66 first reported (obesity).
4. **Hearing loss:** Use of hearing aid, Date H90 first reported (conductive and sensorineural hearing loss), Speech reception threshold (SRT) estimate (left & right).
5. **Smoking**: Pack of years smoking, Current tobacco smoking.
6. **Depression**: Date F32 first reported (depressive episode), Date F33 first reported (recurrent depressive disorder).
7. **Physical inactivity**: Frequency of stair climbing in last 4 weeks, Frequency of walking for pleasure in last 4 weeks, Number of days/week of moderate physical activity (10+ minutes), Number of days/week of vigorous physical activity (10+ minutes), Usual walking pace.
8. **Social isolation:** Social activities, Social isolation, loneliness.
9. **Diabetes:** Date E10 first reported (insulin-dependent diabetes mellitus)
10. **Excessive alcohol consumption:** Alcohol intake versus 10 years previously, Amount of alcohol drunk on a typical drinking day.
11. **Air Pollution:** Particulate matter air pollution (pm2.5); 2010.

(b) (2)

- **High BMI**: BMI.
- **High fasting plasma glucose:** Glycated haemoglobin (HbA1c) (3).
- **Smoking**: Pack of years smoking, Current tobacco smoking.
- **High intake of sugar-sweetened beverages**: Intake of sugar added to coffee, Intake of sugar added to tea

(c) (4)

- **Dementia-related medical histories**: Alzheimer’s disease, all-cause dementia, and vascular dementia.
- **Cognitive Assessments**: Fluid intelligence score.
- **Cholesterol**: Total cholesterol, LDL, HDL
- **White matter hyperintensities**: Total volume of white matter hyperintensities (from T1 and T2_FLAIR images)

#### Target sample for polygenic risk score

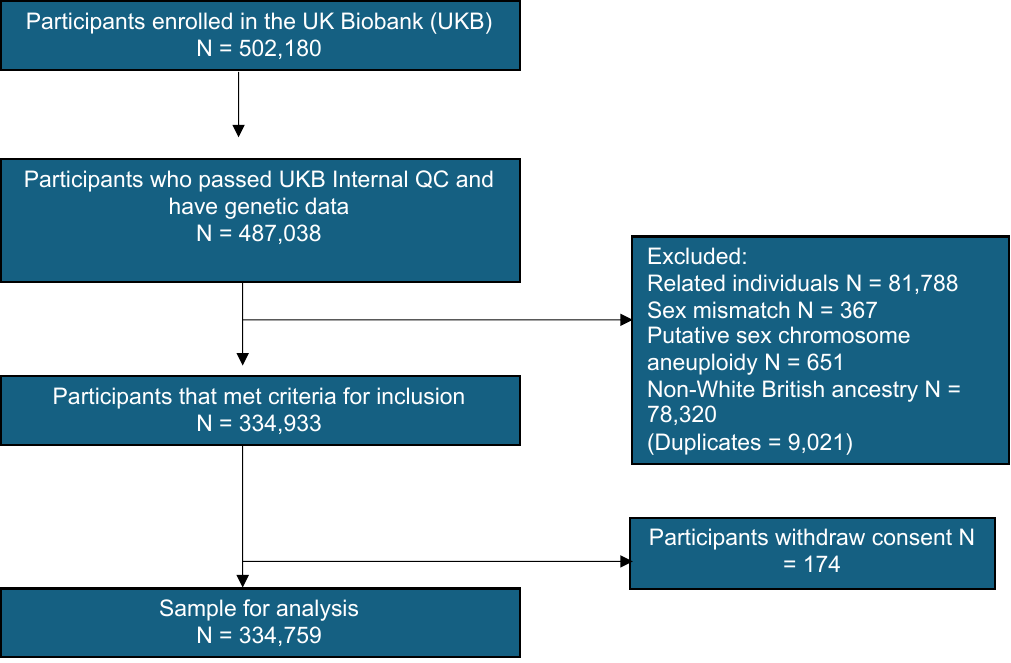
UK Biobank is a population-based study of 503,325 people recruited between 2006 and 2010 from across Great Britain (5,6). The genetic and phenotypic information utilized in this research was accessed from the UK Biobank under application ID: 123335. In Supplementary Figure 2, the flowchart shows the number of participants removed at each stage of the quality control pipeline.

###### Supplementary Figure 1. The flowchart of the UK Biobank study participant selection.

#### Vascular Dementia Meta-Analysis

We conducted a meta-analysis of vascular dementia using the METAL software package. Our analysis integrated summary statistics from two major European genome-wide association studies. The first dataset came from the MEGAVCID consortium (2024), comprising 3,892 cases and 466,606 controls from 11 cohorts with vascular dementia cases (primarily identified through ICD coding systems) and 4 additional control-only cohorts. The second dataset was from the FinnGen project, which included 3,116 cases and 433,066 controls. FinnGen is a comprehensive Finnish genomics initiative analyzing over 500,000 biobank samples in conjunction with health registry data, identifying cases through the "F5_VASCDEM" endpoint based on Finnish health registry records (ICD-10: F01). We performed an inverse-variance weighted meta-analysis to combine these studies, yielding a maximum sample of 7,008 cases and 899,672 controls. Both source studies showed no significant genomic inflation (lambda values within 0.1 of 1), eliminating the need for genomic control correction during meta-analysis.

#### Polygenic Risk Score

We constructed standardised weighted polygenic risk scores (PRS) for VaD for all eligible UK Biobank participants. The VaD PRS incorporated nine independent genetic variants (*P* < 1 × 10^-6^) identified from a mega-analysis of two existing GWAS: (i) the MEGAVCID consortium (N cases: 3,892, N controls: 466,606) (7) and (ii) FinnGen (N cases: 3,116, N controls: 433,066). The VaD PRS was calculated for each participant by multiplying the allele count of each genetic variant by its respective weight in the GWAS and summing across all variants.

#### PHEnome Scan ANalysis Tool (PHESANT)

We conducted the phenome-wide association study (PheWAS) using PHESANT, an open-source R package specifically designed for high-throughput association testing in large-scale biobank data (9). PHESANT implements an automated rule-based method that processes phenotypes according to their specified encoding (continuous, ordered categorical, unordered categorical, or binary), applying appropriate transformations and statistical tests based on variable type. For continuous variables, PHESANT applies an inverse normal rank transformation to approximate normality and mitigate the influence of outliers. For ordered categorical variables, the package fits ordinal logistic regression models, while binary and unordered categorical variables are analysed using logistic and multinomial logistic regression, respectively. The package automatically handles UK Biobank data field encoding, facilitating standardized analysis across thousands of phenotypes while maintaining appropriate statistical methodology for each variable type. All models were adjusted for relevant covariates as described above. In total, 64 UK Biobank variables excluded from PHESANT include (Field ID): assessment (54); polymorphic fields containing values with mixed data (82,92); genetic data description fields (22000, 22001, 22003-6, 22009-22013, 22018-19, 22021, 22027, 22051-2); sex (31), age-related fields (34. 52, 21003, 21022, 21200); assessment centre environment fields (20012-4, 3059, 3065, 3081, 4268, 4275, 4281, 4287, 5149, 5152, 5155, 5164, 6024, 6074-5); and categorical fields with more than one value recorded per person (4232, 4243, 4259, 5090-1, 5136, 5138-5148, 6312, 401, 402, 10691).

#### Follow-up using MR

Of the 558 phenotypes identified in the PheWAS for vascular dementia, and of the 21 previously implicated risk factors of dementia, we followed up 186 phenotypes using two-sample bidirectional Mendelian randomization. We did not follow up remainder phenotypes identified in the PheWAS because of low prevalence (less than 500 cases for binary phenotypes), lack of available genetic instruments, or if they indicated own diagnosis or family history of Alzheimer's disease. For medication-related phenotypes, we used the traits targeted by the medications (e.g., LDL for statins) rather than the medications themselves.

Our primary analytical method was inverse variance weighted (IVW) regression, which provides the most precise estimates when all genetic variants are valid instruments. However, since IVW assumes no horizontal pleiotropy (where genetic variants affect the outcome through pathways independent of the exposure), we also used MR-Egger regression, which can detect and account for directional pleiotropy through its intercept test, sacrificing precision for increased robustness (29), and the weighted median estimator, which provides consistent estimates even when up to 50% of the information comes from invalid instruments (30).

**Sensitivity analyses**

However, since IVW assumes no horizontal pleiotropy (where genetic variants affect the outcome through pathways independent of the exposure), we also used MR-Egger regression, which can detect and account for directional pleiotropy through its intercept test, sacrificing precision for increased robustness (29), and the weighted median estimator, which provides consistent estimates even when up to 50% of the information comes from invalid instruments (30).

### Supplementary Results

#### Polygenic Risk Score

##### Vascular dementia polygenic risk score

To examine the relationship between VaD PRS and various neurological and dementia-related outcomes, we conducted association analyses across multiple exposures. A standard deviation (SD) increase in the VaD PRS was significantly associated with an increased risk of vascular dementia, Alzheimer’s disease, and all-cause dementia, with the strongest association observed for Alzheimer’s disease (AD) (OR= 1.86, 95% CI: 1.81 to 1.91, R^2^=0.047). A significant association was found between VaD PRS and white matter hyperintensity (WMH) volume in the oldest tertile, after adjusting for total intracranial volume (ICV) and assessment centre (β = 0.025, 95% CI: 0.011 to 0.040, R^2^=0.0003). Our sensitivity analysis included genome-wide significant SNPs (P < 5 x 10^-8^; N = 4) in the VaD PRS, which showed similar significant associations with dementia subtypes and WMH volume in the oldest tertile: AD (OR= 1.94, 95% CI: 1.88 to 1.99, R^2^=0.053), and adjusted WMH (β = 0.030, 95% CI: 0.015 to 0.044, R^2^=0.0004).

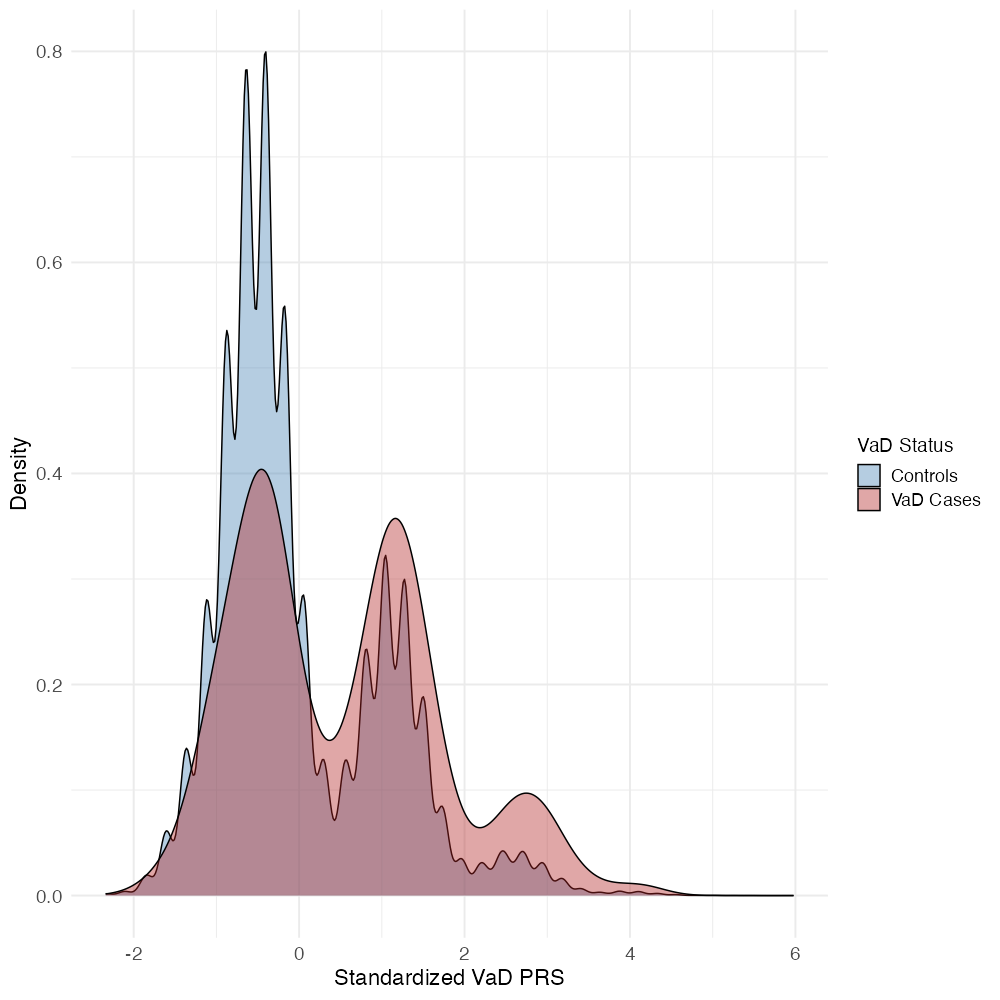

###### Supplementary Figure 2. Density curves of standardized vascular dementia polygenic risk scores (PRS) stratified by case-control status.

#### Phenome-wide Association Study

##### Association of vascular dementia polygenic risk score and the phenome

###### Age 62-72 years:

The oldest age tertile included 16,300 tests ranked by P value. We identified 558 associations below a P value threshold corresponding to a 5% FDR (*P value* threshold =1.69x10^-3^). Of these, 328 results had a P value lower than a stringent Bonferroni corrected threshold of 3.07x10^-6^ (**Supplementary Table 6**).

###### Age 53-62 years:

The middle age tertile included 15,698 tests ranked by P value. We identified 440 associations below a P value threshold corresponding to a 5% FDR (P value threshold =1.37x10^-3^). Of these, 265 results had a P value lower than a stringent Bonferroni corrected threshold of 3.19x10^-6^ (**Supplementary Table 7**).

###### Age 39-53 years:

The youngest age tertile included 15,698 tests ranked by P value. We identified 440 associations below a P value threshold corresponding to a 5% FDR (P value threshold =1.37x10^-3^. Of these, 265 results had a P value lower than a stringent Bonferroni corrected threshold of 3.19x10^-6^ (**Supplementary Table 8**).

###### Entire Sample:

The entire sample included 21,266 tests ranked by P value. We identified 701 associations below a P value threshold corresponding to a 5% FDR (P value threshold =1.64x10^-3^. Of these, 405 results had a P value lower than a stringent Bonferroni corrected threshold of 2.35x10^-6^ (**Supplementary Table 9**).

##### Association of vascular dementia polygenic risk score with only genome-wide significant hits and the phenome

###### Age 62-72 years:

The oldest age tertile included 16,580 tests ranked by P value. We identified 848 associations below a P value threshold corresponding to a 5% FDR (*P value* threshold =2.55x10^-3^). Of these, 336 results had a P value lower than a stringent Bonferroni corrected threshold of 3.02x10^-6^ (**Supplementary Table x**).

###### Age 53-62 years:

The middle age tertile included 9,215 tests ranked by P value. We identified 448 associations below a P value threshold corresponding to a 5% FDR (P value threshold =2.41x10^-3^). Of these, 258 results had a P value lower than a stringent Bonferroni corrected threshold of 5.43x10^-6^ (**Supplementary Table x**).

###### Age 39-53 years:

The youngest age tertile included 8,857 tests ranked by P value. We identified 371 associations below a P value threshold corresponding to a 5% FDR (P value threshold =1.99x10^-3^. Of these, 225 results had a P value lower than a stringent Bonferroni corrected threshold of 5.65x10^-6^ (**Supplementary Table x**).

##### Sensitivity Analysis:

We assessed pleiotropy using heterogeneity statistics and pleiotropy-robust methods (supplementary table 13). Several instrumental variables demonstrated highly heterogeneous effects on VaD risk (Q statistic *P* < 0.05), suggesting potential pleiotropic effects. These included genetic instruments for blood biomarkers (blood assay measures, high fasting plasma glucose), anthropometric and metabolic traits (arm and trunk mass, basal metabolic rate), cardiovascular traits (coronary heart disease, diastolic and systolic blood pressure), and neurological and lifestyle factors (years of smoking, white matter hyperintensities). The MR-Egger intercept of apolipoprotein A (OR = 1.02, 95% CI: 1.01-1.03, P = 0.002) and C-reactive protein (OR = 1.02, 95% CI: 1.01-1.03, P = 0.001) showed evidence of directional pleiotropy.

To address potential bias from sample overlap between exposure and outcome datasets, we implemented MR-LAP analyses. For vascular dementia, MR-LAP revealed that generally very little bias was detected due to overlapping samples, likely due to using strong instruments (11). Where bias was detected, conventional IVW effect sizes were likely underestimated (e.g. for schizophrenia, depression, gastroesophageal reflux disease, white matter hyperintensities, BMI, social activities (e.g., religious group), oily fish intake, raw vegetable intake, and gamma-glutamyl transferase (Supplementary Table 15). Notably, MR-LAP identified evidence of causal effects for multiple phenotypes that did not show evidence in the conventional IVW analyses (neurological and cognitive factors (Alzheimer's disease and multiple cognitive function tests involving trail making, digit substitution, and reaction time measures; metabolic and body composition factors such as vitamin D levels, arm fat percentage, and alanine aminotransferase; and lifestyle factors including moderate and vigorous physical activity, degree of fatty acid unsaturation, and non-oily fish intake, supplemental table 16).

### Supplementary Figures

#### Vascular Dementia PheWAS Plots

Legends in each graph indicate age tertiles and entire sample. Effect estimates represent an SD change in the phenotype per 1 unit increase in the standardized polygenic risk score for vascular dementia. Error bars represent 95% confidence intervals. Each tertile consists of 111,656 participants and the exact sample size for each phenotype are in the Supplementary Data file. Continuous effect estimates were derived from linear regression models and are in standard deviations, while binary/ordered effect estimates were derived from binary logistic models and effect estimates are on the log odds scale.

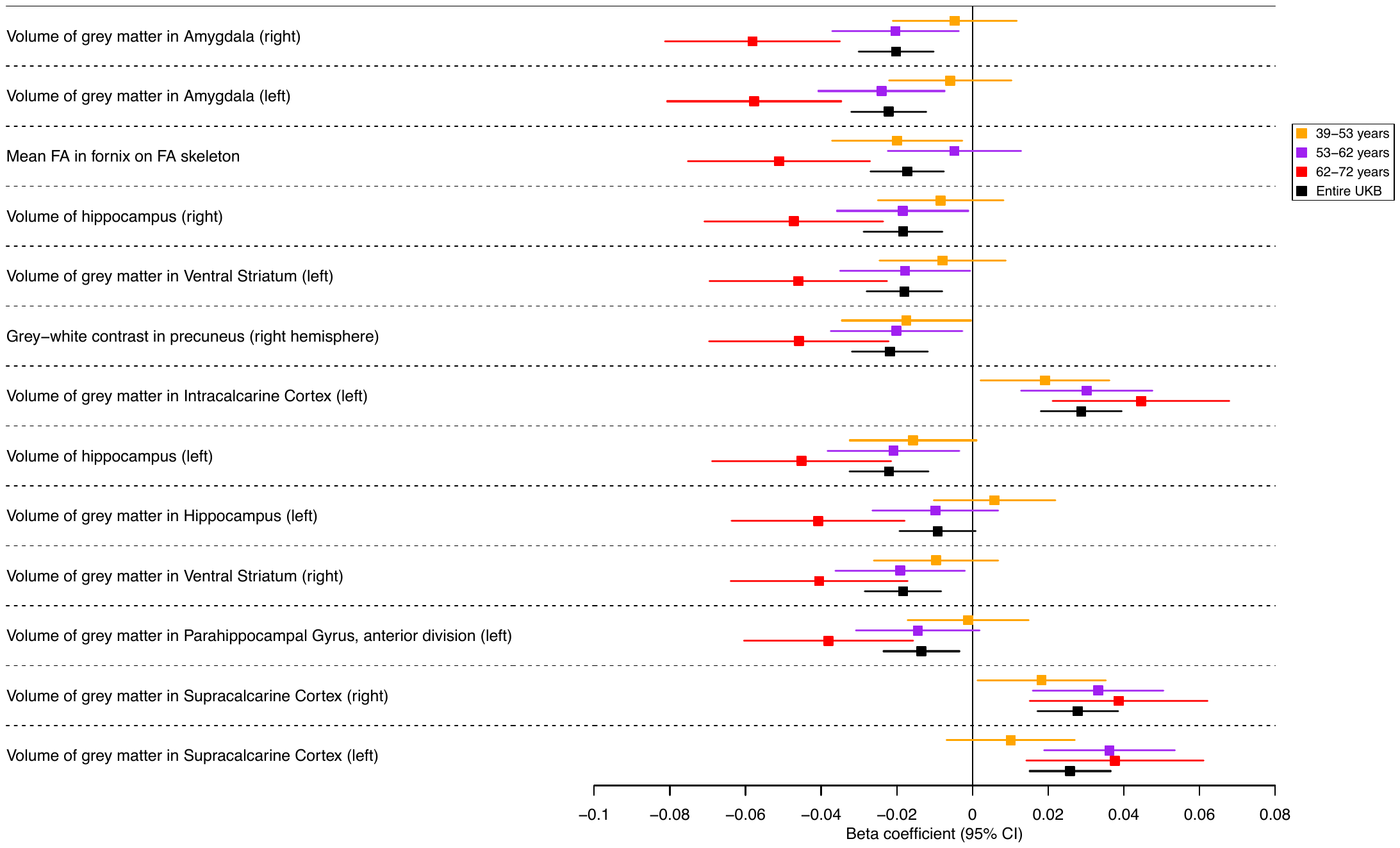

###### Supplementary Figure 3. Forest plots showing effect estimates for the association between vascular dementia polygenic risk score and brain imaging phenotypes, **by age tertile**.

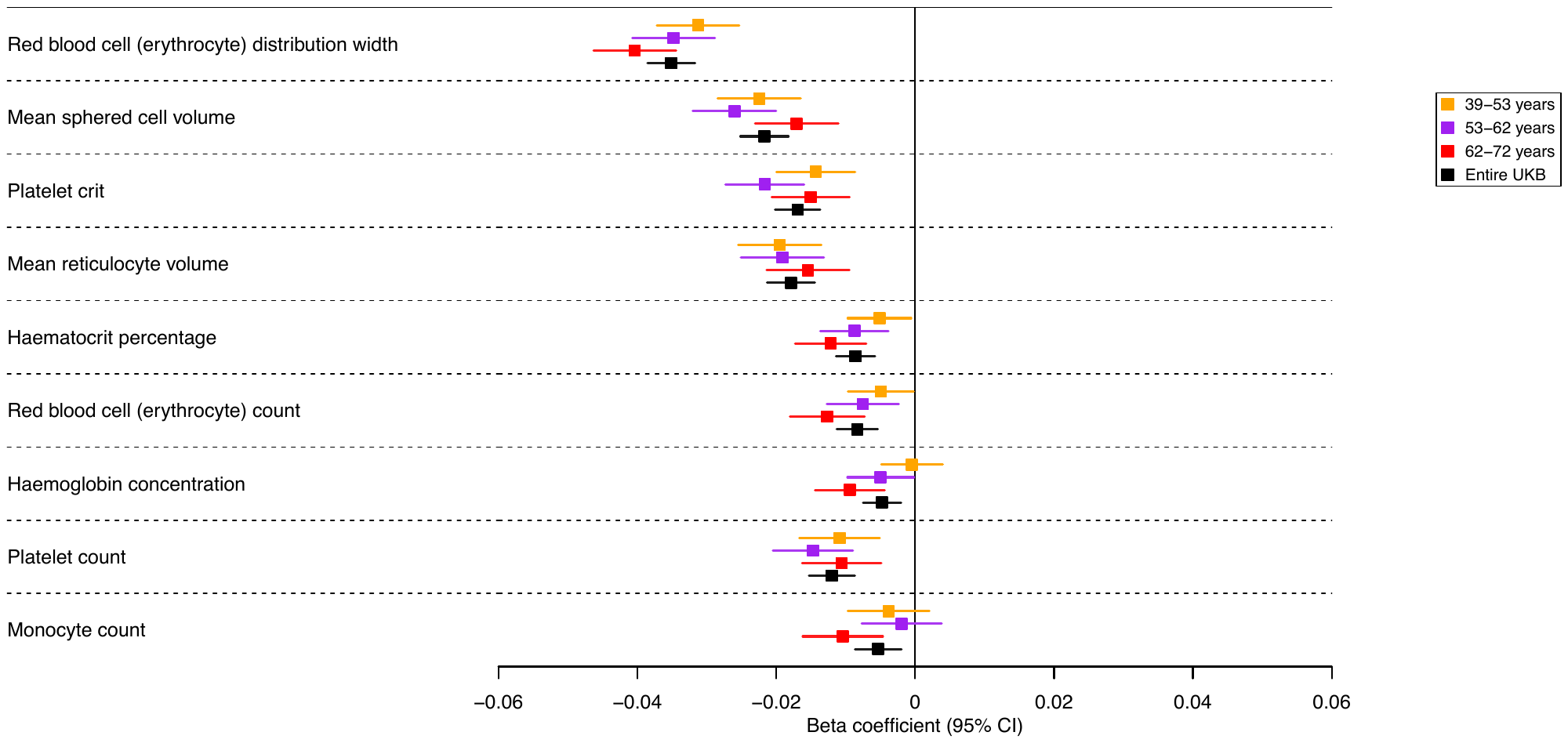

###### Supplementary Figure 4. Forest plots showing effect estimates for the association between vascular dementia polygenic risk score and biological measures from red blood cell measures, by age tertile.

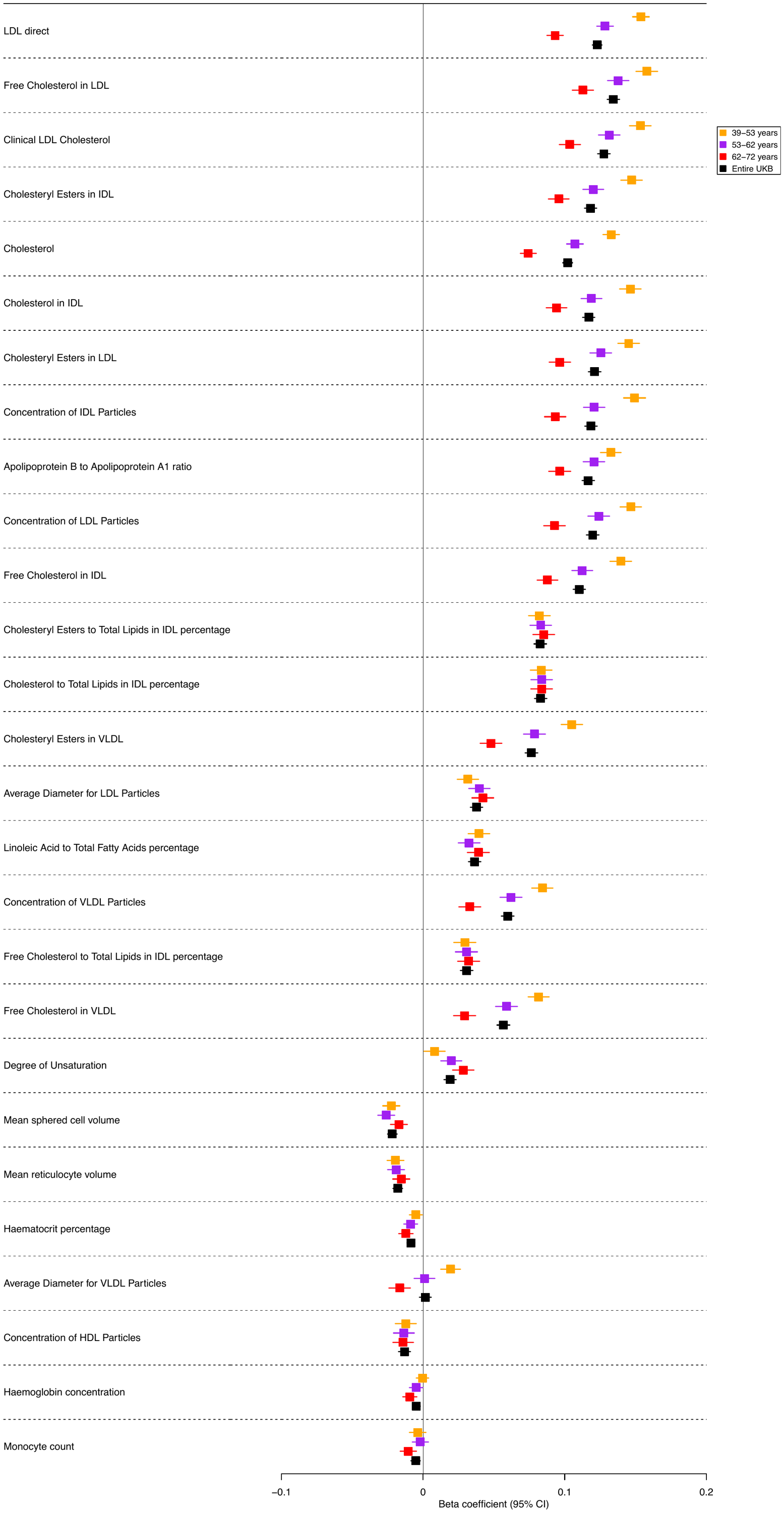

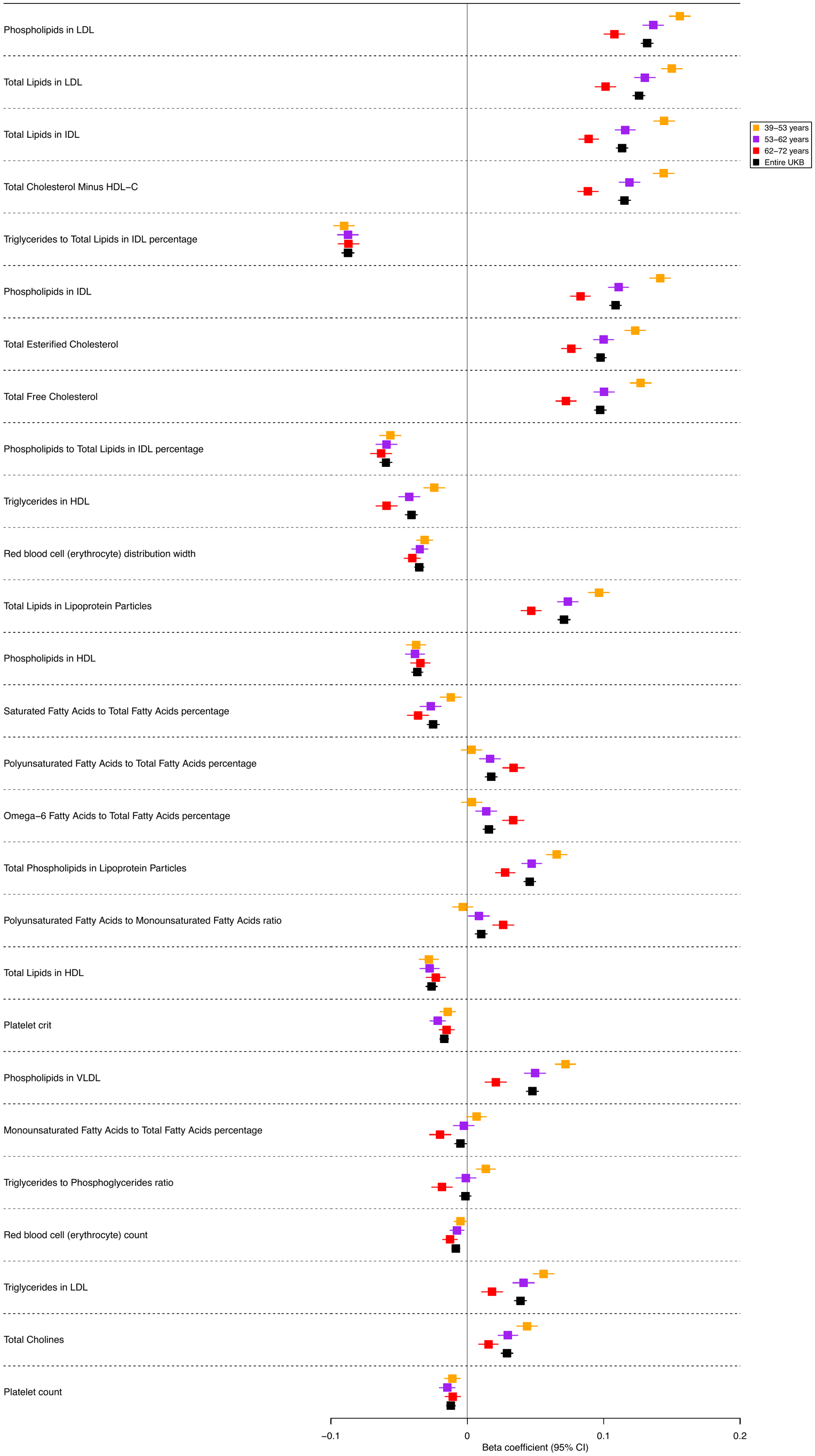

###### Supplementary Figure 5. Forest plots showing effect estimates for the association between vascular dementia polygenic risk score and biological measures from blood assay results**,** by age tertile**.**

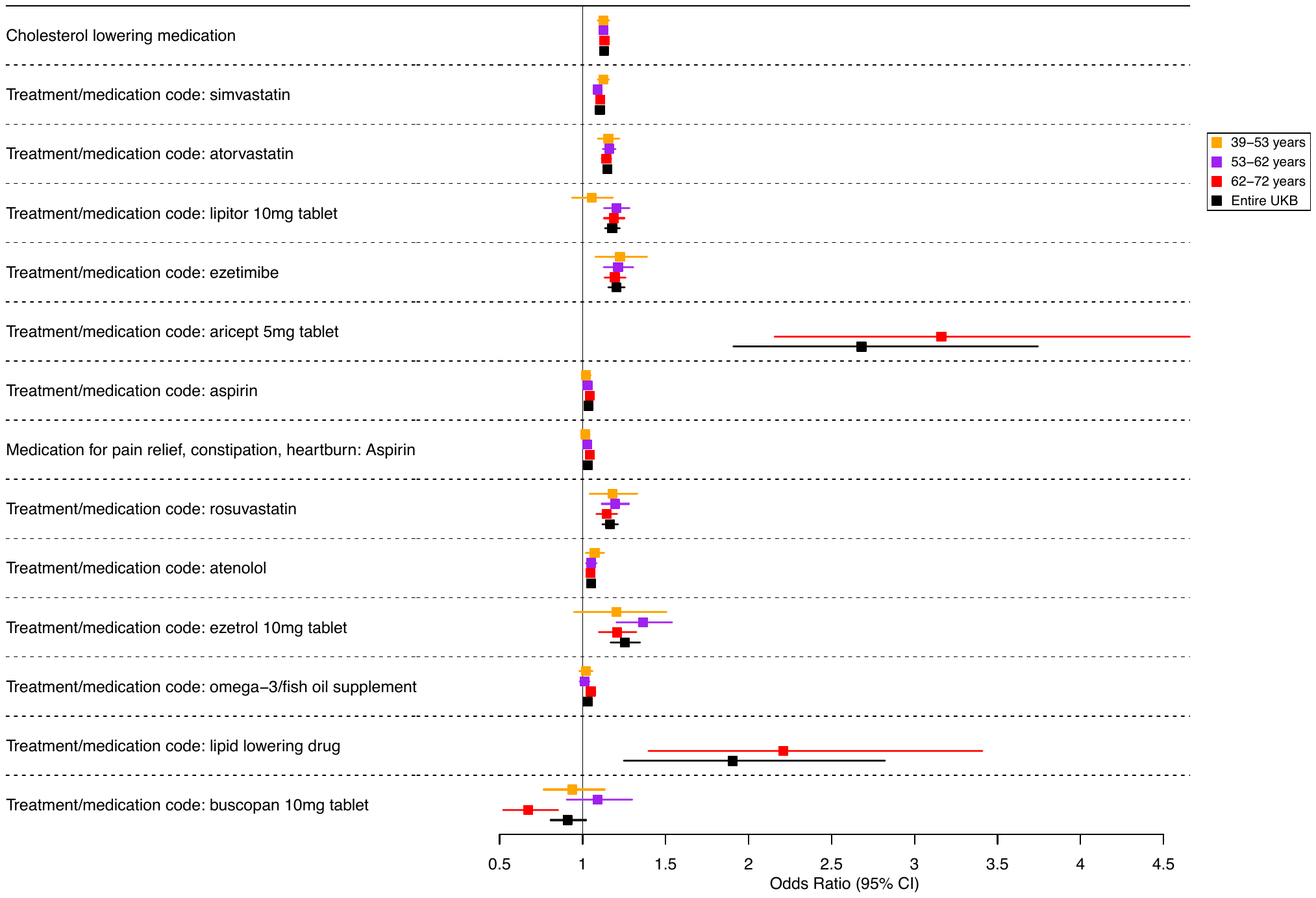

###### Supplementary Figure 6. Forest plots showing effect estimates for the association between vascular dementia polygenic risk score and medication use**,** by age tertile.

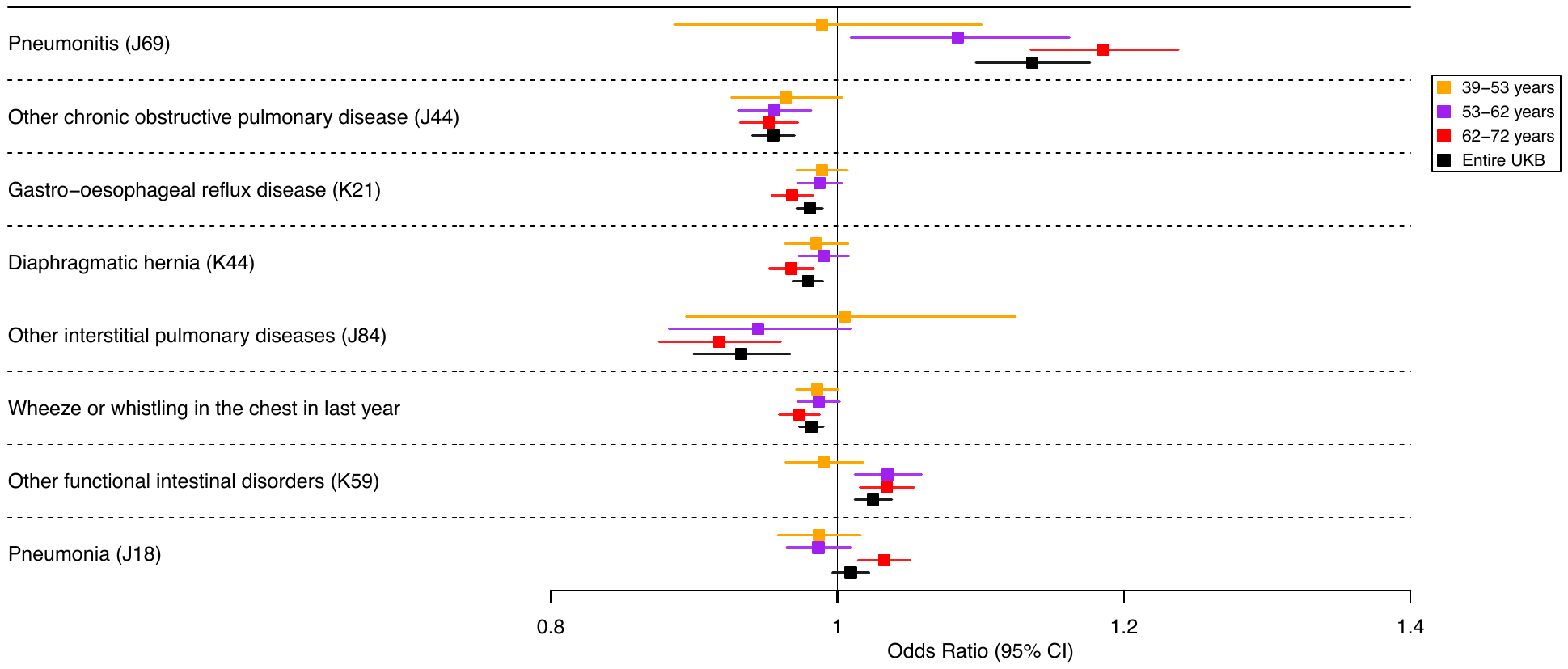

###### Supplementary Figure 7. Forest plots showing effect estimates for the association between vascular dementia polygenic risk score and respiratory & gastrointestinal medical history, by age tertile.

###### Additional Results:

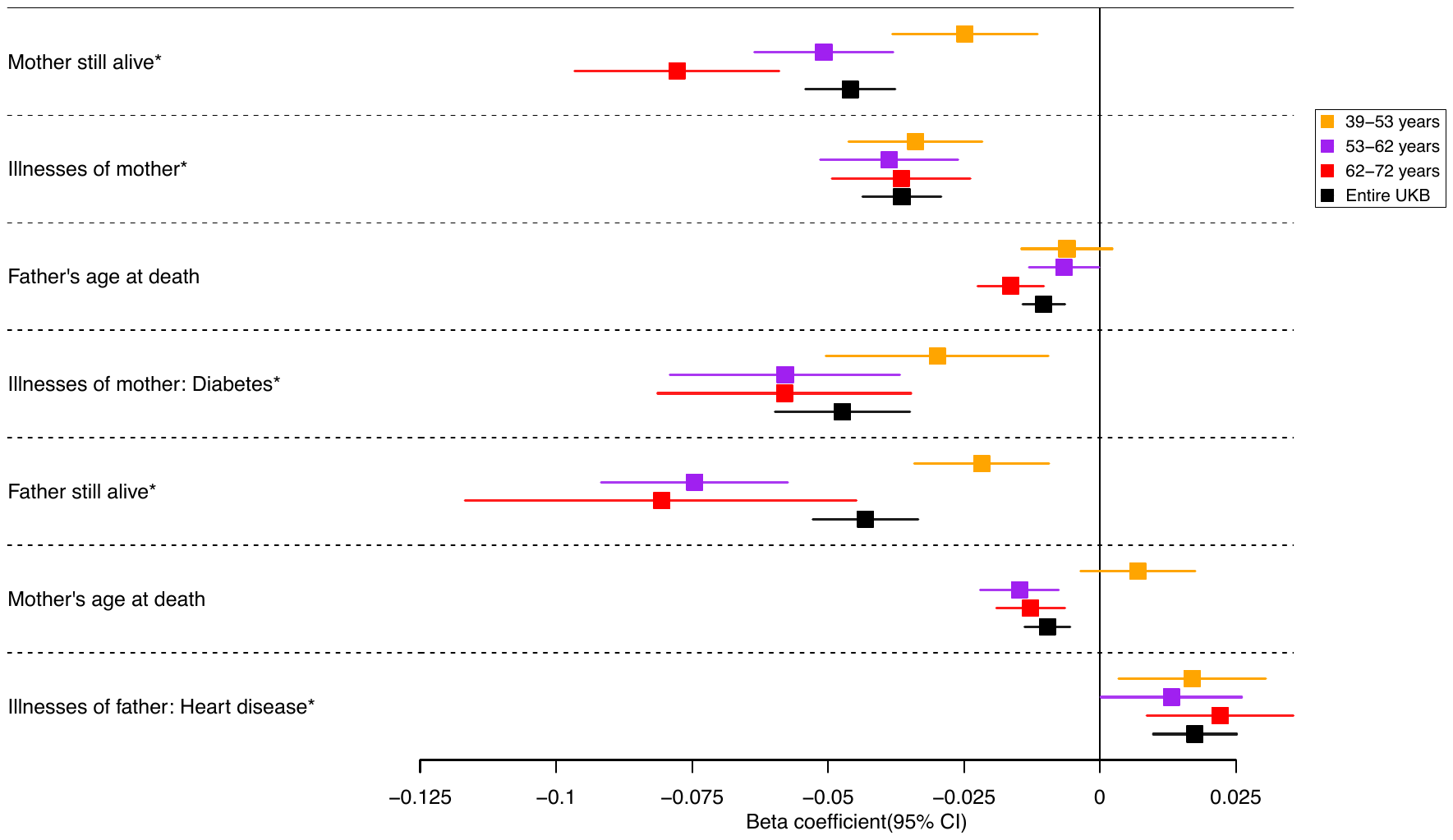

###### Supplementary Figure 8. Forest plots showing effect estimates for the association between vascular dementia polygenic risk score and family history, by age tertile. Legends in each graph indicate age tertiles and entire sample * Effect estimates were derived from binary logistic models; effect estimates are on the log odds scale.

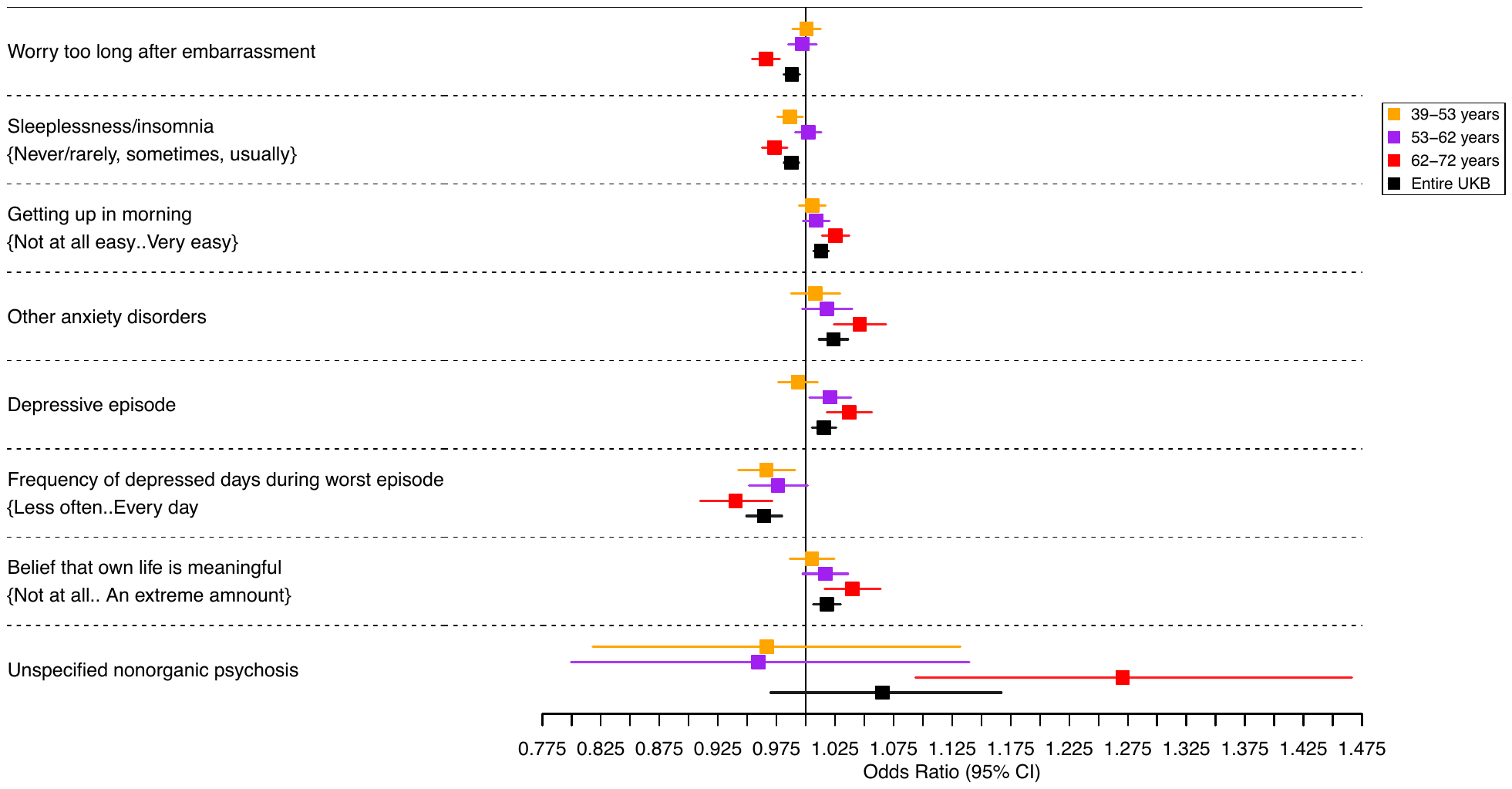

###### Supplementary Figure 9. Forest plots showing effect estimates for the association between vascular dementia polygenic risk score and psychiatric disorders, by age tertile.

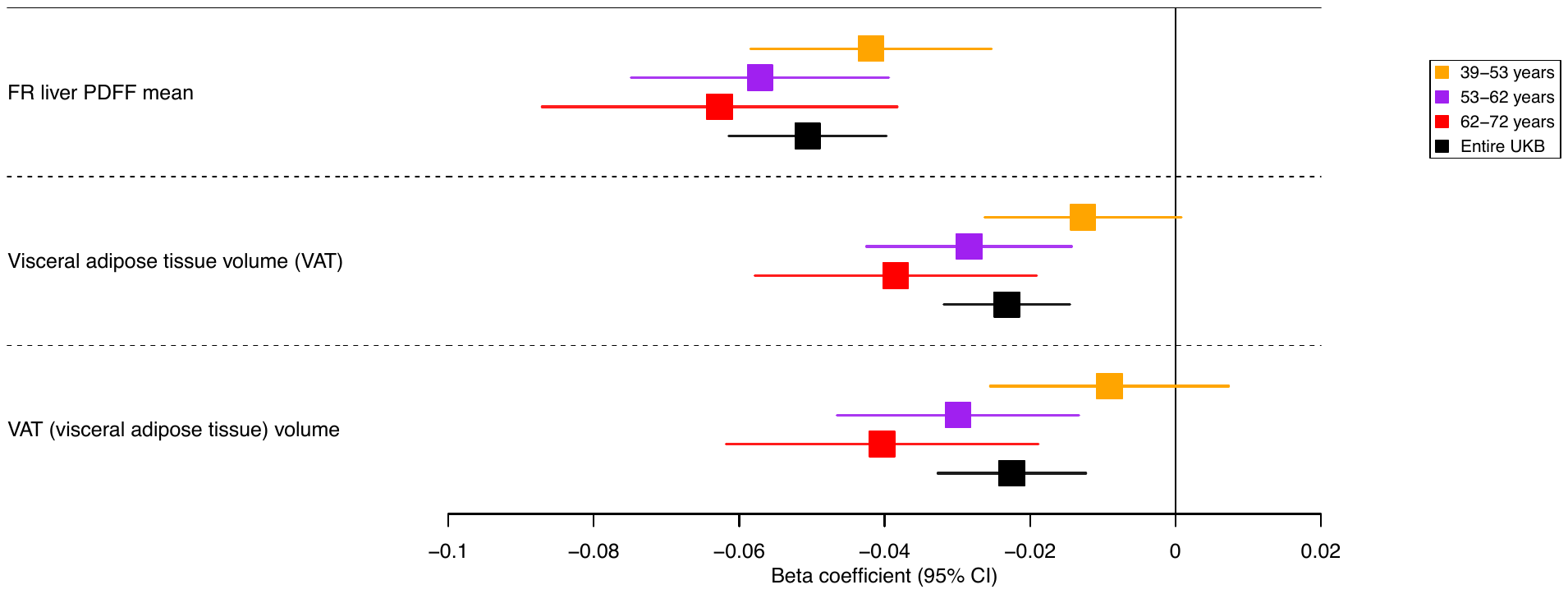

###### Supplementary Figure 10. Forest plots showing effect estimates for the association between vascular dementia polygenic risk score and imaging phenotypes, by age tertile.

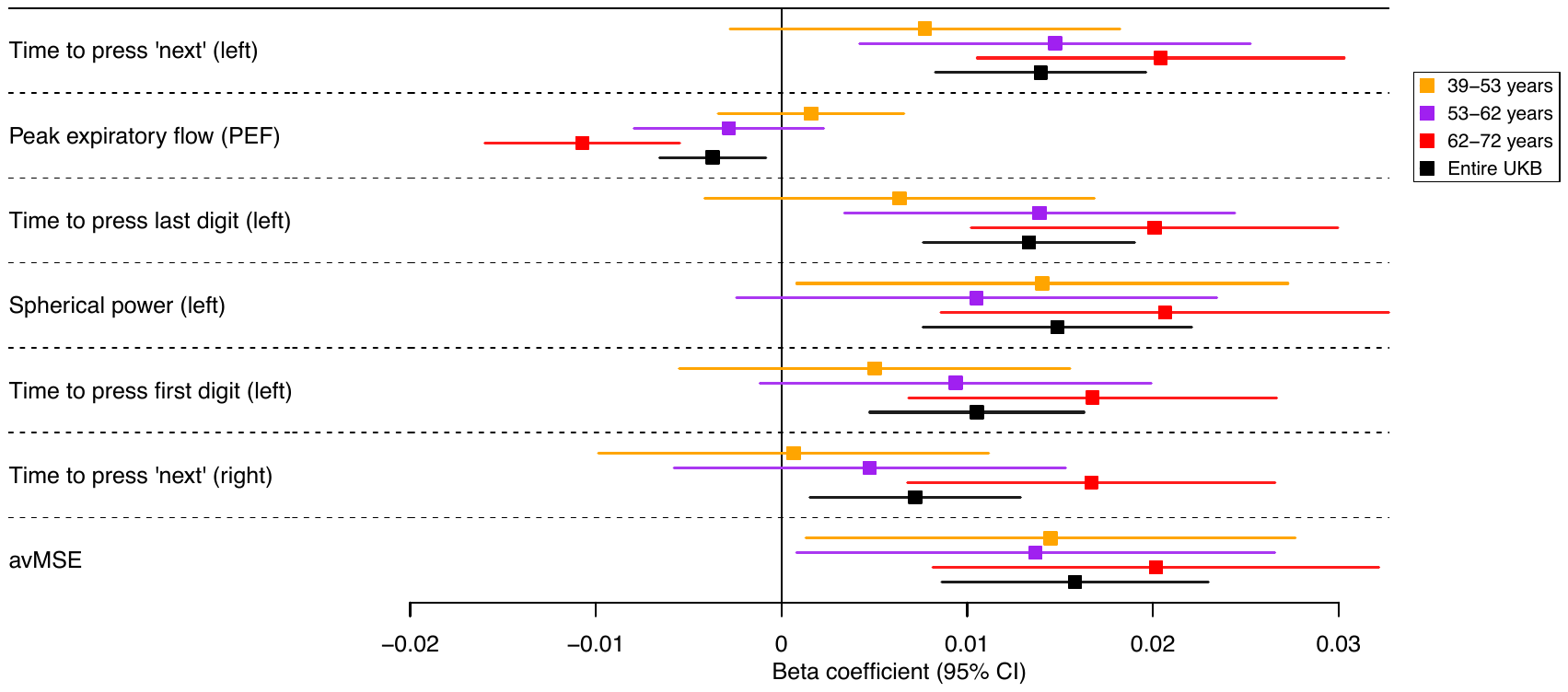

###### Supplementary Figure 11. Forest plots showing effect estimates for the association between vascular dementia polygenic risk score and cardiovascular, respiratory, vision, and hearing function, **by age tertile**.

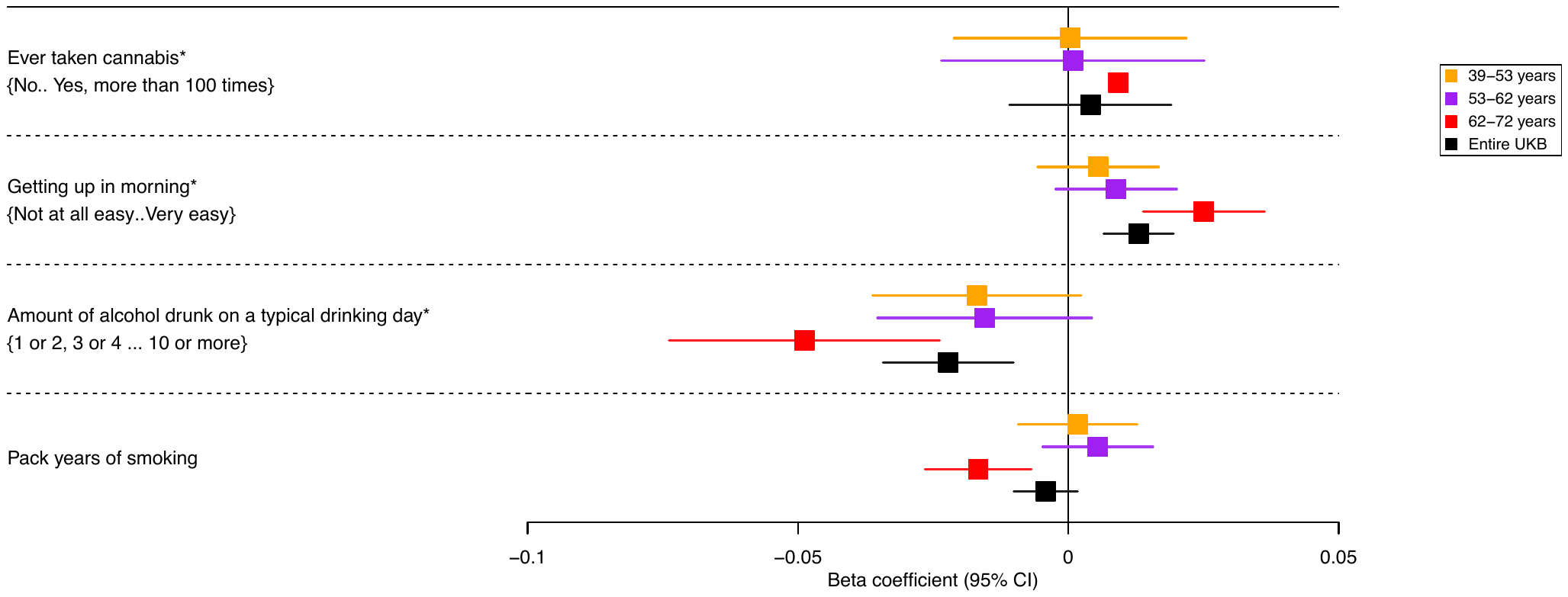

###### Supplementary Figure 12. Forest plots showing effect estimates for the association between vascular dementia polygenic risk score and lifestyle factors, by age tertile. *Effect estimates were derived from binary logistic/ordered-logistic regression models, and effect estimates are on the log odds scale.

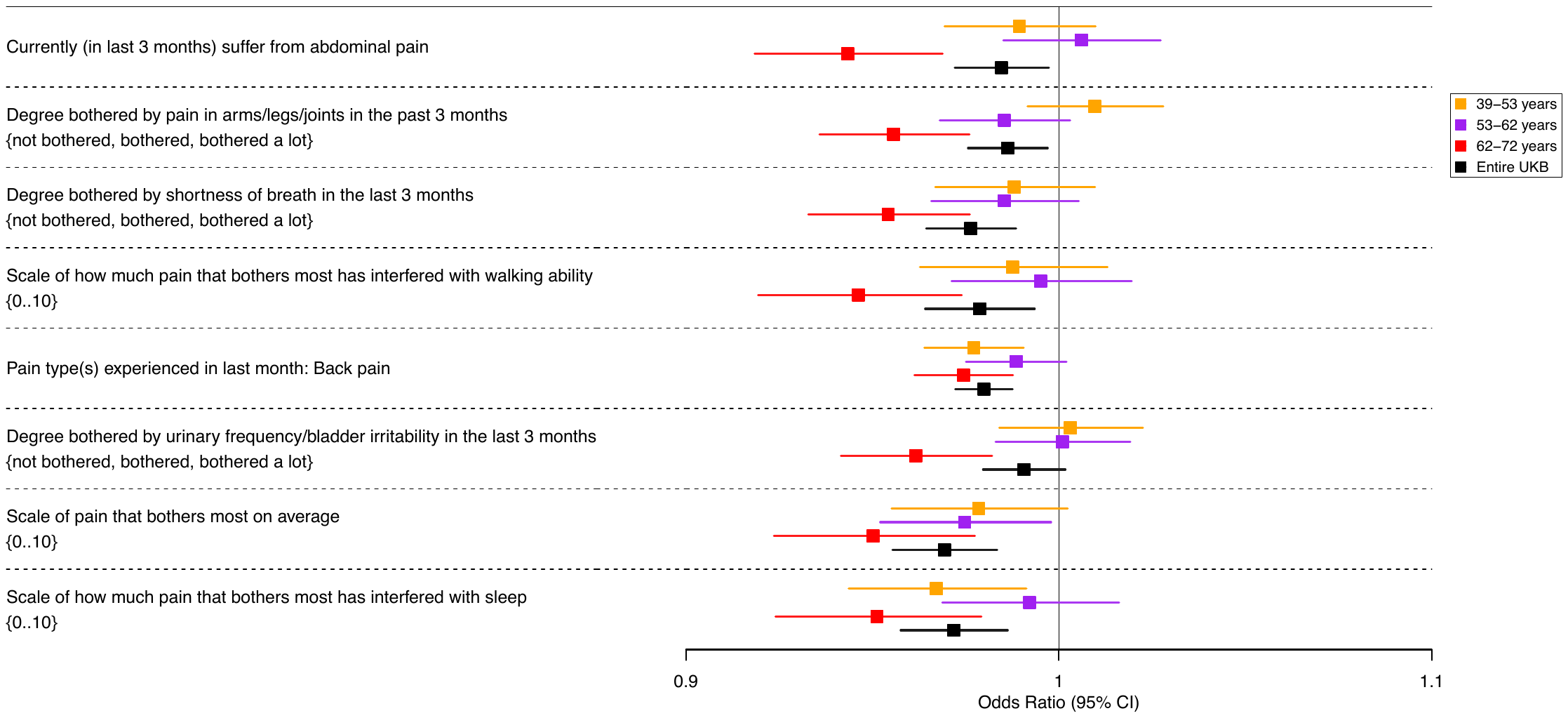

###### Supplementary Figure 13. Forest plots showing effect estimates for the association between vascular dementia polygenic risk score and the experience of pain, by age tertile.

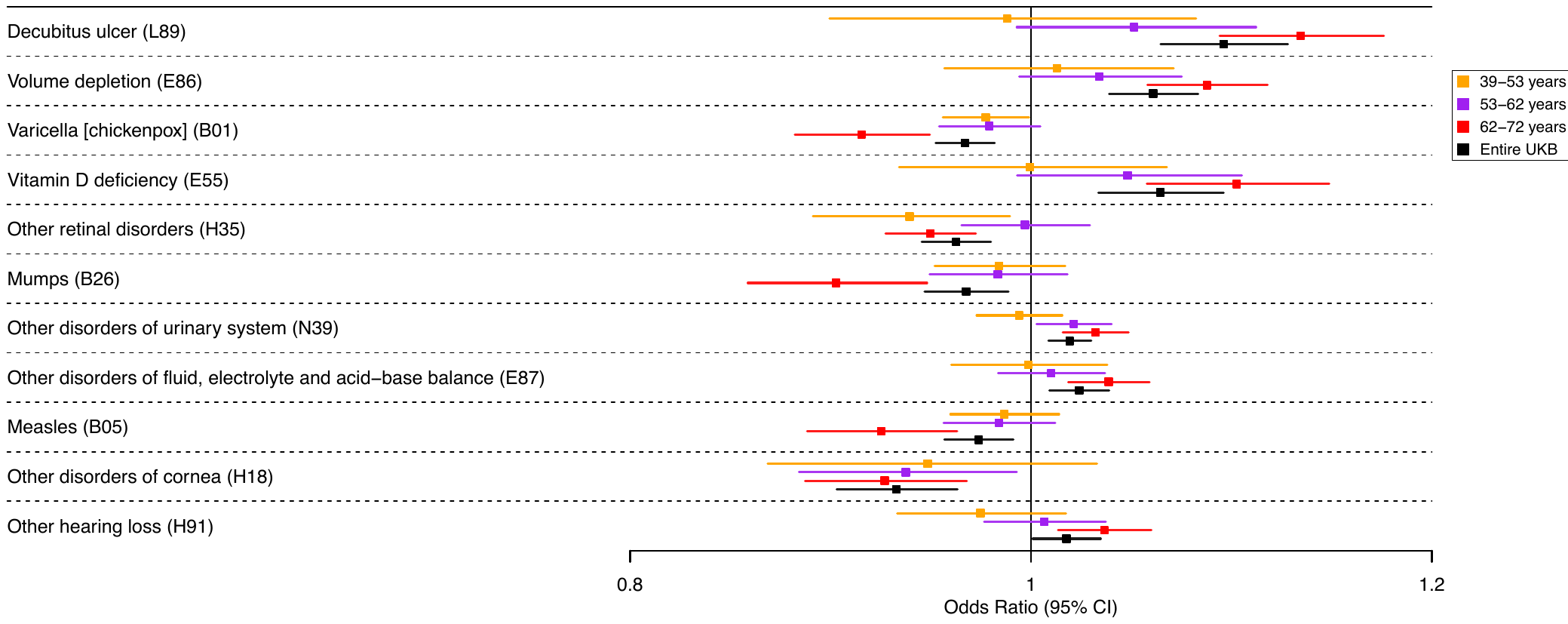

###### Supplementary Figure 14. Forest plots showing effect estimates for the association between vascular dementia polygenic risk score and infectious & other medical history, by age tertile.

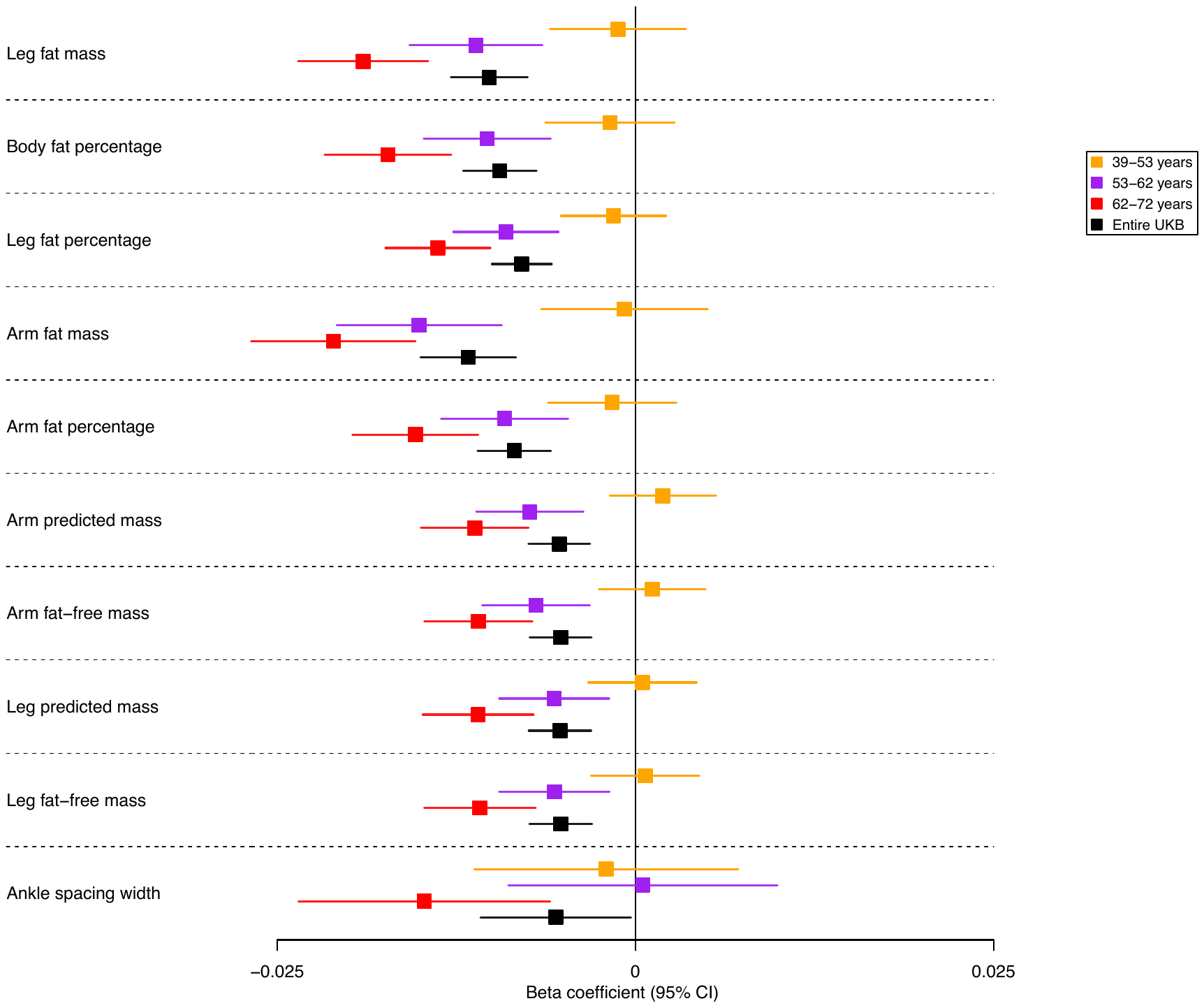

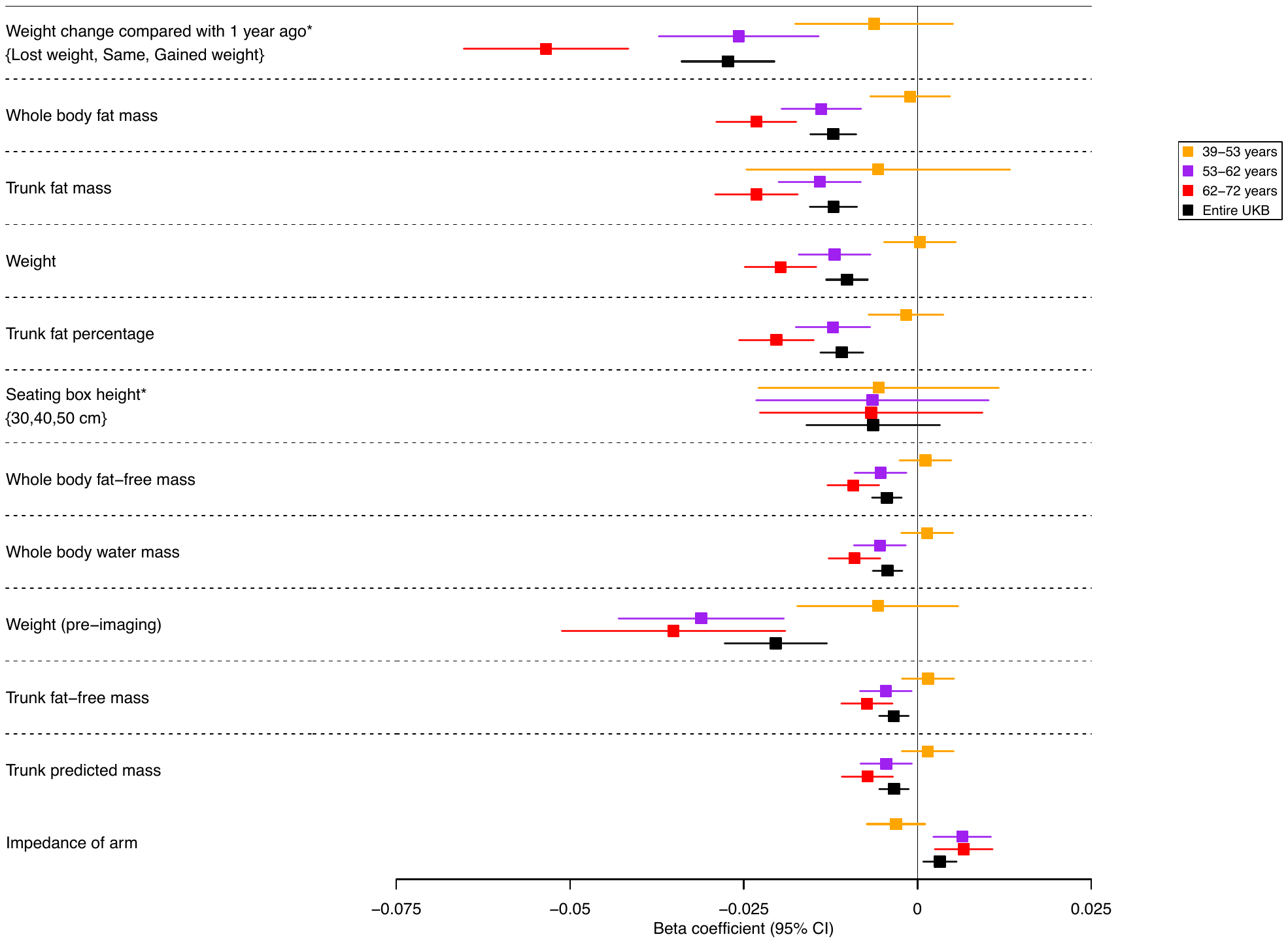

###### Supplementary Figure 15. Forest plots showing effect estimates for the association between vascular dementia polygenic risk score and anthropometric and body composition, by age tertile. *Effect estimates were derived from binary logistic/ordered-logistic regression models, and effect estimates are on the log odds scale.

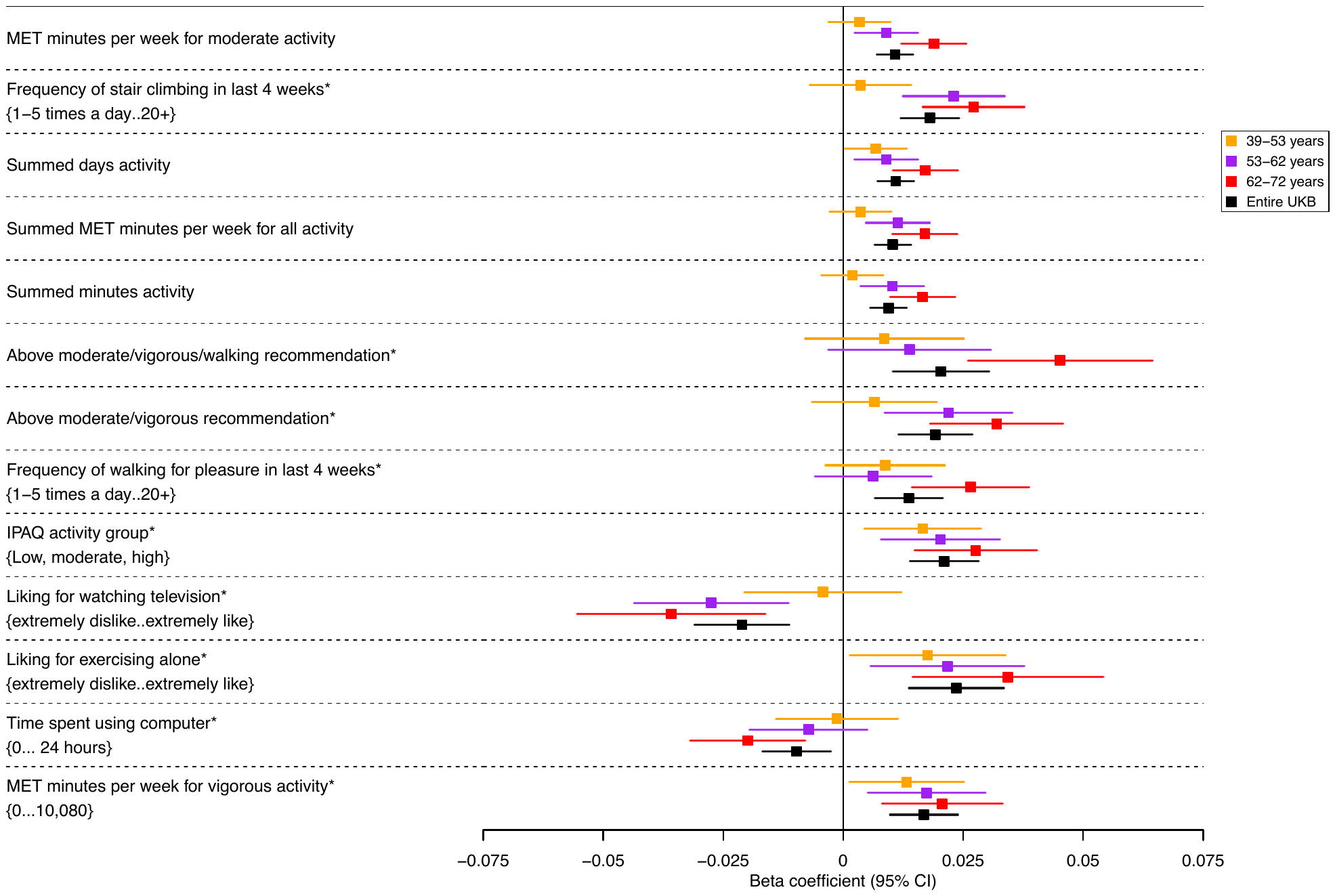

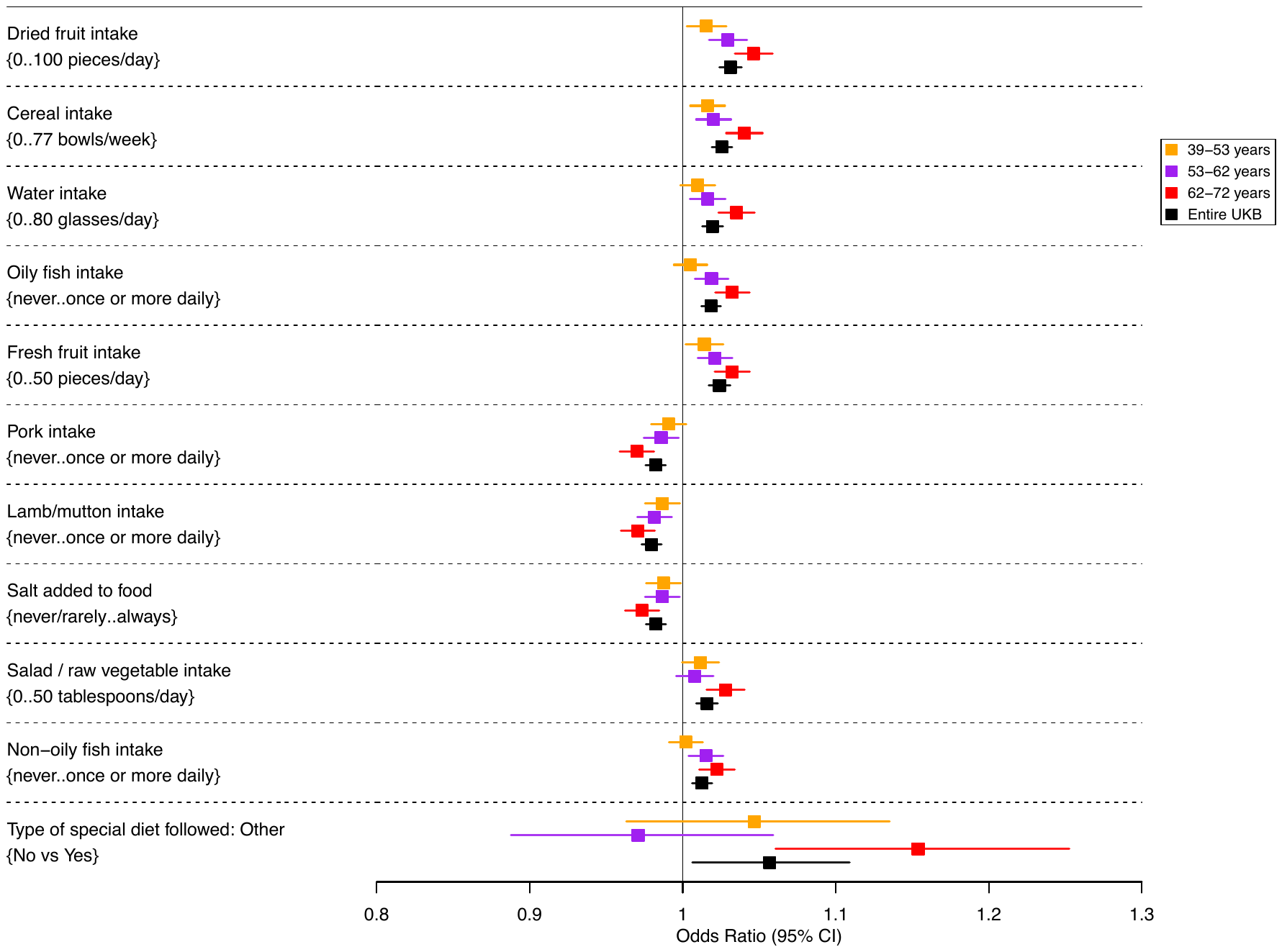

###### Supplementary Figure 16. Forest plots showing effect estimates for the association between vascular dementia polygenic risk score with physical activity measures and dietary factors, by age tertile. *Effect estimates were derived from binary logistic/ordered-logistic regression models, and effect estimates are on the log odds scale. For dietary measures the effect estimates were derived from binary logistic models, effect estimates are on the log odds scale.

##### Sensitivity Analysis: Comparison Between Original PheWAS and Adjusted PheWAS

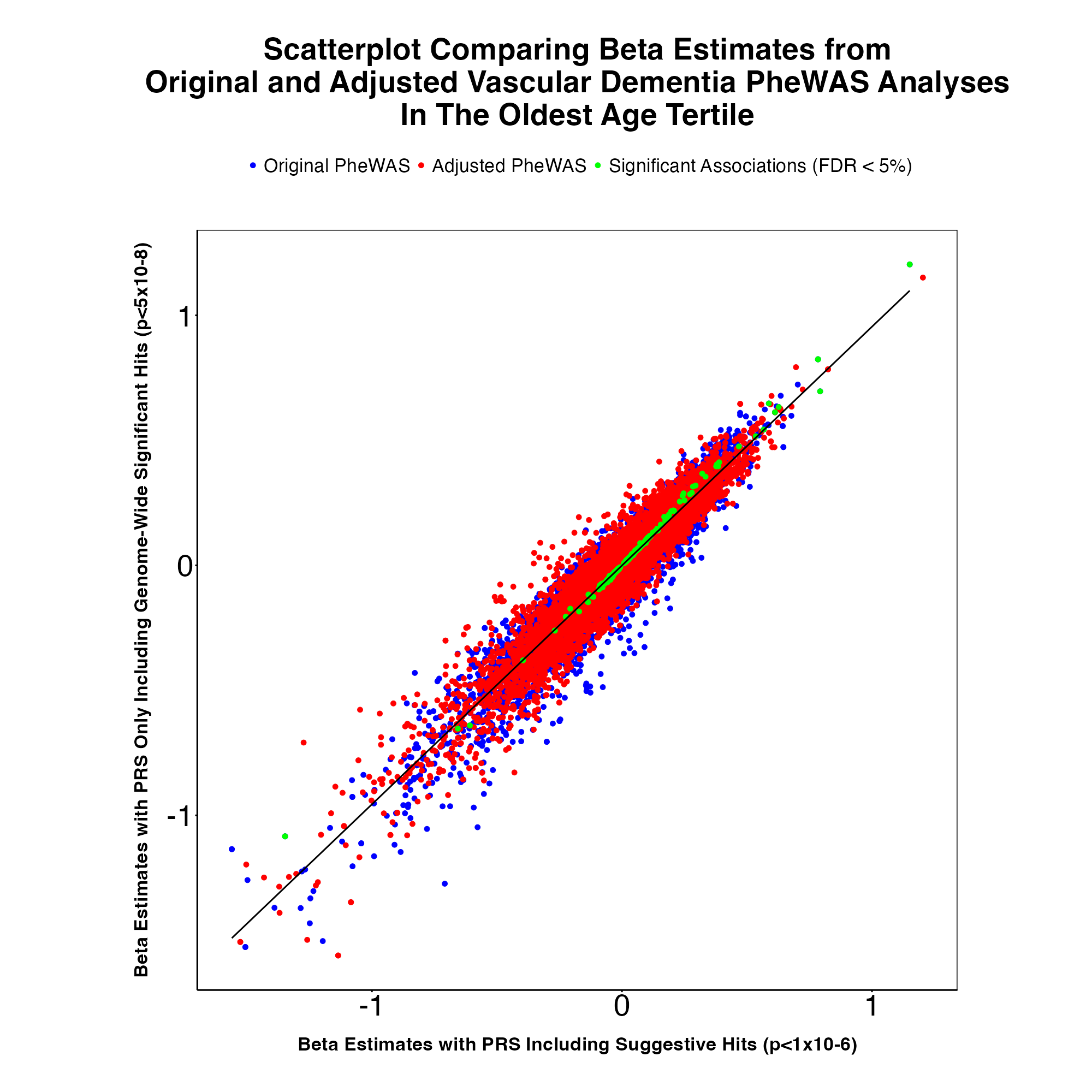

###### Supplementary Figure 17. Scatterplot comparing all beta estimates from the original and adjusted vascular PheWAS analyses in the oldest tertile group. Plot compares beta estimates from the original PheWAS (blue dots), which included suggestive (p < 1x10^-6^) SNPs, and the adjusted PheWAS (red dots) where we only included genome-wide significant (p < 5x10^-8^) SNPs in our PRS. Significant associations (false discovery rate (FDR) < 5%) in the adjusted analysis are highlighted in green.

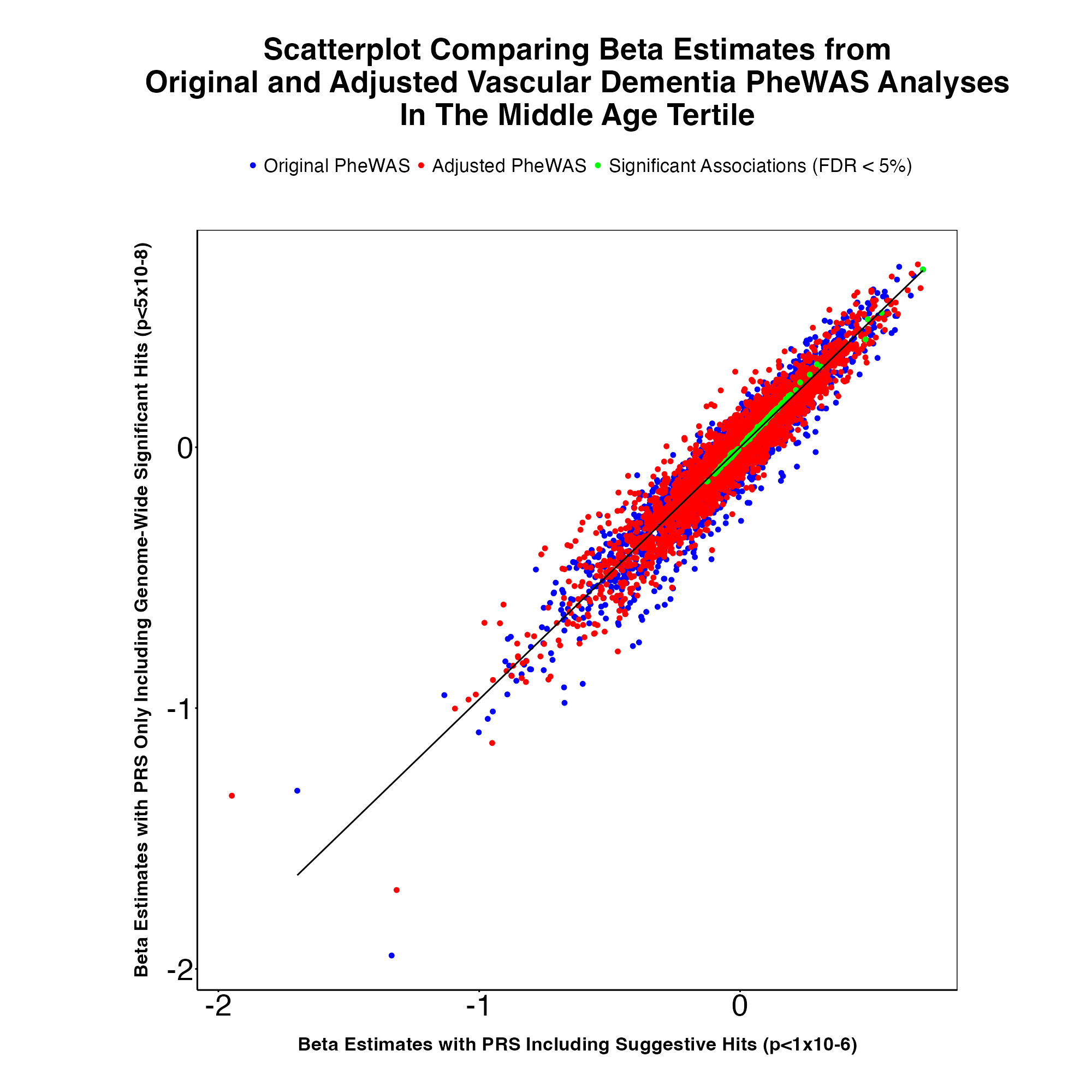

###### Supplementary Figure 18. Scatterplot comparing all beta estimates from the original and adjusted vascular PheWAS analyses in the middle tertile group. Plot compares beta estimates from the original PheWAS (blue dots), which included suggestive (p < 1x10^-6^) SNPs, and the adjusted PheWAS (red dots) where we only included genome-wide significant (p < 5x10^-8^) SNPs in our PRS. Significant associations (false discovery rate (FDR) < 5%) in the adjusted analysis are highlighted in green.

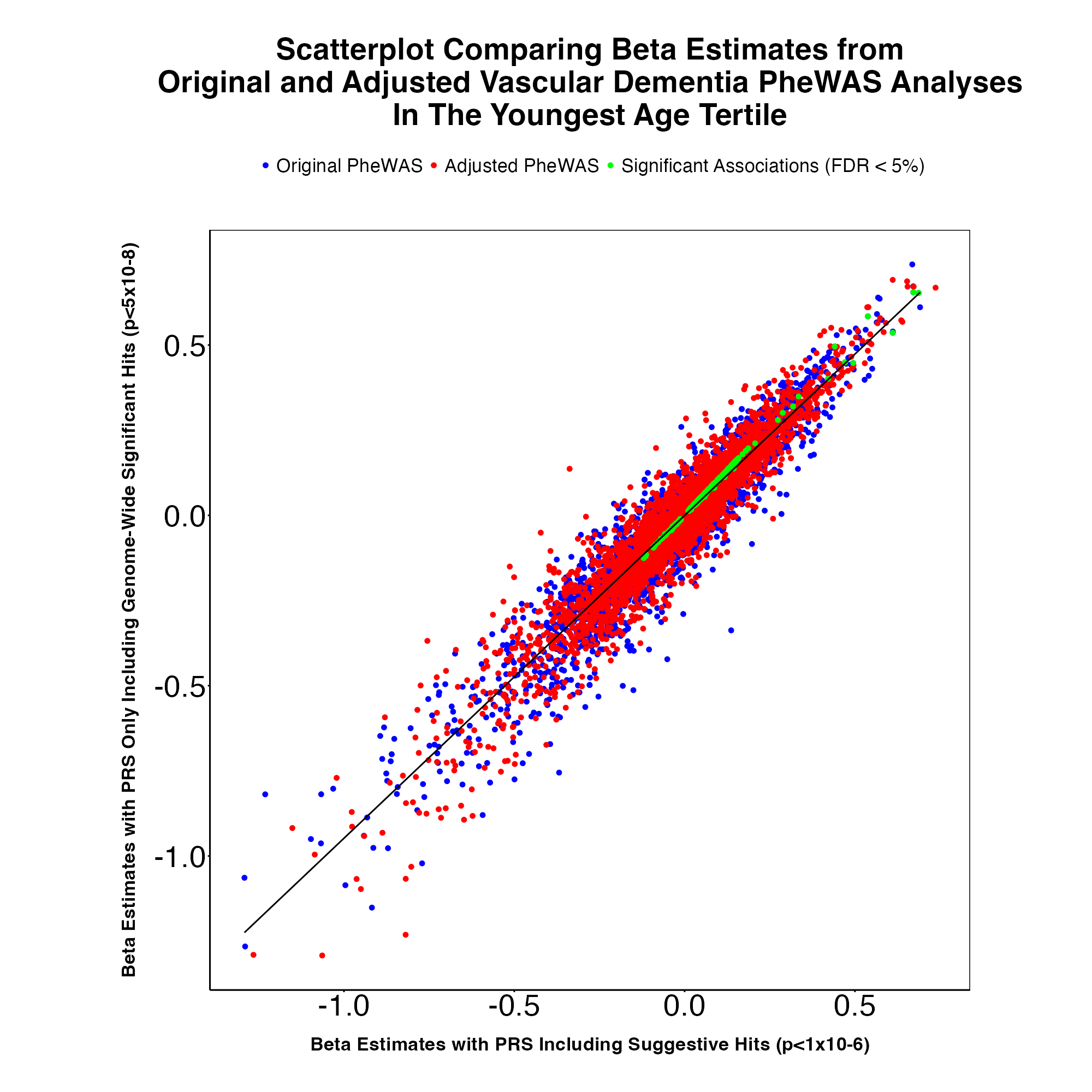

###### Supplementary Figure 19. Scatterplot comparing all beta estimates from the original and adjusted vascular PheWAS analyses in the youngest tertile group. Plot compares beta estimates from the original PheWAS (blue dots), which included suggestive (p < 1x10^-6^) SNPs, and the adjusted PheWAS (red dots) where we only included genome-wide significant (p < 5x10^-8^) SNPs in our PRS. Significant associations (false discovery rate (FDR) < 5%) in the adjusted analysis are highlighted in green

#### Mendelian Randomization Output

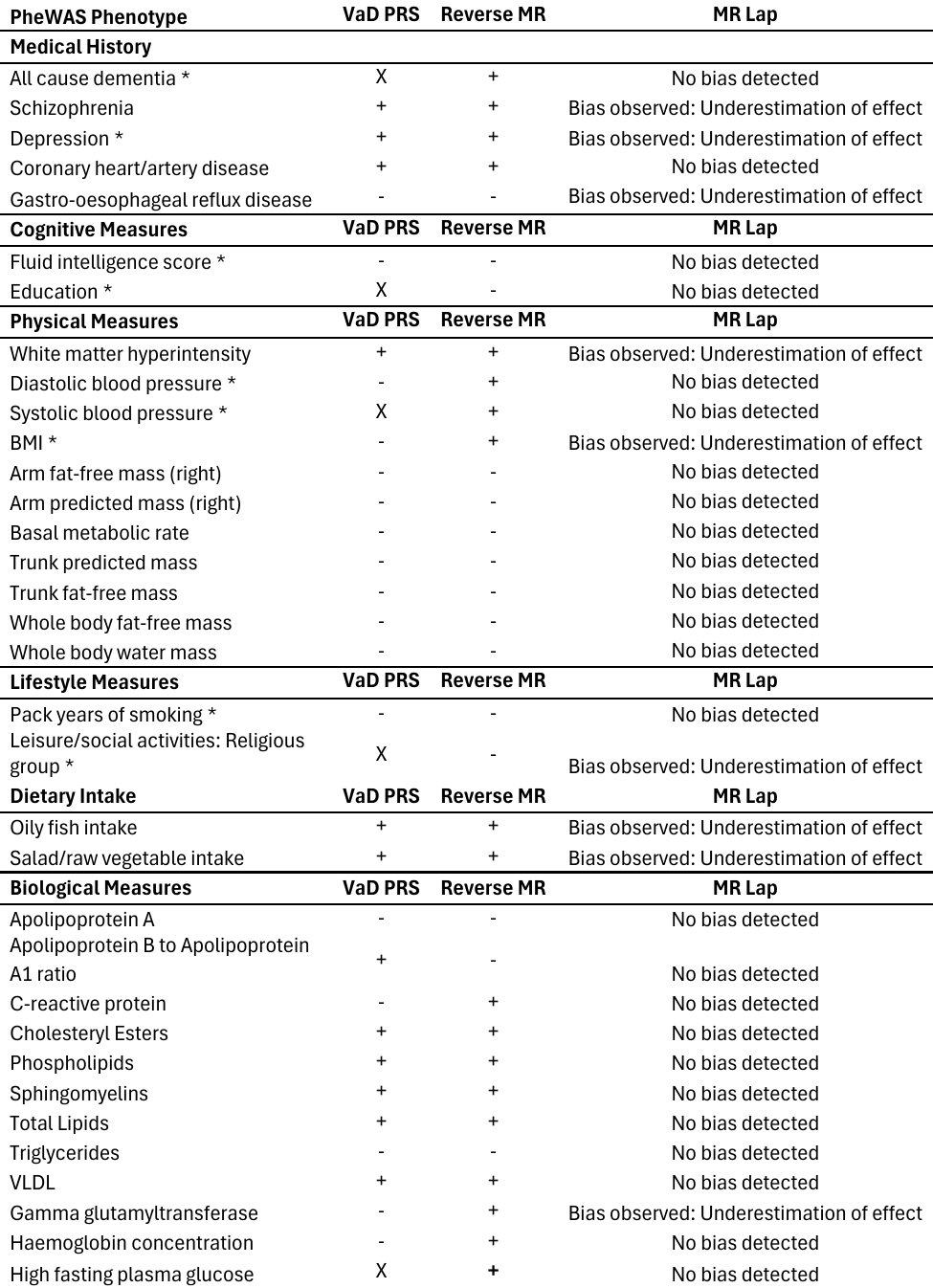

###### Supplementary Figure 20. Significant and suggestive association of vascular dementia polygenic risk score with the phenome, and estimated effect of each phenotype using Mendelian randomization. These findings showed evidence of association in the MR framework, following a correction for multiple testing using a strategy controlling for the false discovery rate. + and – indicate the direction of the coefficient for phenotypes associated with vascular dementia using two-sample MR. X represents associations that were consistent with the null. *Previously implicated risk factors of all-cause dementia.
